# Diverse and weakly immunogenic *var* gene expression facilitates malaria infection

**DOI:** 10.1101/2023.12.27.23300577

**Authors:** Inayat Bhardwaj, Prince B. Nyarko, Asrar Ba Ashn, Camille Cohen, Sukai Ceesay, Jane Achan, Edgard Dabira, Rike Nakajima, Aarti Jain, Omid Taghavian, Algis Jasinskas, Philip L. Felgner, Umberto D’Alessandro, Teun Bousema, Mark Travassos, Ovidiu Radulescu, Antoine Claessens

**Affiliations:** LPHI University of Montpellier and CNRS, Montpellier, France; Malaria Research Program, Center for Vaccine Development and Global Health, University of Maryland School of Medicine, Baltimore, United States of America; Medical Research Council Unit The Gambia at the London School of Hygiene and Tropical Medicine, Banjul, The Gambia; Department of Medical Microbiology and Radboud Center for Infectious Diseases, Radboud University Medical Center, Nijmegen, The Netherlands; Vaccine Research & Development Center, Department of Physiology & Biophysics, School of Medicine, University of California, Irvine, Irvine, California, United States of America

**Author notes:** equal contribution (Inayat Bhardwaj, Prince B. Nyarko).

## Abstract

*Plasmodium falciparum* is believed to escape immunity via antigenic variation, mediated in part by 60 *var* genes. These genes undergo mutually exclusive expression and encode the PfEMP1 surface antigen. The frequency of *var* switching and the immunogenicity of each expressed PfEMP1 remain unclear. To this end, we carried out a Controlled Human Malaria Infection (CHMI) study with 19 adult African volunteers in The Gambia to gain insight into the effect of naturally acquired immunity on the expressed *var* gene repertoire during early phase of an infection. Our findings demonstrated a strong correlation between the diversity of *var* expression, quantified through entropy, and infection outcome. Low-immunity individuals were characterised by high *var* entropy profiles, higher parasitaemia, and lower sero-recognised PfEMP1 domains compared to high-immunity individuals. For the first time we recorded the probability of *var* gene switching *in vitro* and of turnover *in vivo*, enabling us to estimate both intrinsic switching and negative-selection effects. These processes are rapid, resulting in estimated turnover/switching probabilities of 69% - 97% and 7% - 57% per generation, *in vivo* and *in vitro*, respectively. *Var* (PfEMP1) expression triggered time-dependent humoral immune responses in low immunity individuals, with many PfEMP1 domains remaining weakly immunogenic. We conclude that the role of intrinsic *var* switching is to reset and maintain a diverse *var* repertoire. The high *var* switching rates and weak PfEMP1 immunogenicity benefit parasite survival during the CHMI.

## 1 Introduction

While all human-infecting *Plasmodium* species invade, grow and replicate within erythrocytes, *P. falciparum* is distinct in its ability to modify the surface of infected cells.These changes impact erythrocytes’ cytoadhesive properties, with late-pigmented trophozoite and schizont stages sequestering within the microvasculature. Sequestration is essential for the avoidance of splenic clearance of late-stage infected red blood cells (iRBC), but can result in microvascular obstruction and the release of pro-inflammatory cytokines, which are key features of malaria pathogenesis [63]. Of the parasite Variant Surface Antigens (VSA), Plasmodium falciparum Erythrocyte Membrane Protein 1 (PfEMP1) is the major ligand binding to human endothelial receptors. PfEMP1 is encoded by a family of *∼*60 *var* genes that undergo mutually exclusive expression; meaning, a single type of *var* gene is expressed at each cycle (with peak transcription at 16 hours post invasion). Each *P. falciparum* isolate typically contains a unique set of 60 *var* sequences, making the worldwide pool of *var* gene sequences virtually infinite [7]. Despite this mind-boggling polymorphism, *var* genes are classified into four main sub-families, named Group A, B, C and E, based on their upstream sequence (Ups) and some conserved motifs [47]. Almost all PfEMP1s have a head structure composed of an N-terminal sequence (NTS), followed by a total of four to nine Duffy binding-like (DBL) and cysteine-rich interdomain region (CIDR) domains, and a semi-conserved intra-cellular acidic terminal sequence (ATS) domain. Crucially, this nomenclature has been repeatedly associated with malaria pathogenesis [65]. In younger children and in cerebral malaria cases, parasites tend to express group A *var* genes; more specifically, PfEMP1s containing a CIDR1*α* domain that mediates binding the brain endothelial receptor EPCR. Conversely, Group B and group C PfEMP1 are expressed in uncomplicated malaria cases and bind to endothelial cell receptor CD36. The best-characterised PfEMP1 variant expression and infection prognosis is pregnancy-associated malaria, where *var2csa* (group E) binds chondroitin sulfate A (CSA) in the placenta, leading to the sequestration of infected red blood cells in placental blood vessels [37, 62]. Other polymorphic VSA include *rif* (*∼*180 copies), with a role in dampening anti-malarial immunity [52], and *stevor* (*∼*30 copies), which are key for iRBC stiffness [39].

Owing to their extracellular exposure, cytoadherent molecules on the surface of infected erythrocytes are also the primary antigenic targets of the immune system, eliciting variant-specific antibody responses. Although anti-RIFIN [22] and anti-STEVOR [40] antibodies have been shown to be functional for promoting immune effector mechanisms, PfEMP1 is thought to be the main target of both total and functional anti-VSA antibodies [12]. Antibodies against group A PfEMP1s are quickly acquired in life and show moderate level of strain-transcending cross-reactivity 21, likely providing protection against the most severe forms of malaria. On the other hand, immunity against Group B and C PfEMP1 takes years, if ever, to develop. Theoretical modeling and experimental data predicts immune acquisition against PfEMP1 variants likely leads to sequential and/or homogeneous *var* expression - a phenomenon postulated to maintain infection chronicity by restricting the number of PfEMP1 variants due to partially cross-reactive and short-lived epitope-specific antibodies [49, 67, 23]. Consequently, in malaria-endemic regions, older children and adults with partially acquired immunity are frequently asymptomatic, i.e. individuals often carry parasite loads without exhibiting symptoms of malaria [35, 19, 13]. *P. falciparum* parasites have thus evolved unique mechanisms of regulating the expression of adhesive surface protein variants to evade the host’s adaptive immunity [17, 18]. The proportions of parasites expressing different *var* genes in a population can change through two mechanisms; intrinsic switching and turnover as a result of selection. Intrinsic switching is the probabilistic change of the expressed gene from one intraerythrocytic cycle to the next [63, 54]. *In vivo*, in addition to intrinsic switching, parasites expressing specific PfEMP1 can be recognized and eliminated by the host immune system. Both mechanisms contribute to the turnover rate, describing how frequently the repertoire of expressed *var* genes change. Low switching and turnover rates are thought to promote prolonged infections by preventing the depletion of the entire gene repertoire [14]. However, as discussed in [14], even the lowest *in vitro* rates estimated from [27] are not capable of explaining very long infections such as chronic malaria. Moreover, it is conceivable that rapid, as opposed to slow switching, provides an advantage for parasites, enabling them to evade recognition by the host immune system during the early stages of an infection. Here, we revisit these concepts and perform an investigation of switching under *in vitro* and turnover under *in vivo* conditions using a Controlled Human Malaria Infections (CHMI) study.

CHMI studies in which human volunteers are infected with *P. falciparum* sporozoites or intra-erythrocytic stages, using the NF54/3D7 clone have highlighted the inherent differences in expression of various *var* genes, with group B being predominantly expressed at the early stages of infection, group A to a lesser extent and group C almost entirely absent [4, 3, 38, 26]. Here, we examined *var* gene transcription at multiple timepoints *in vivo* and *in vitro* in a CHMI study carried out with semi-immune individuals in The Gambia, using the NF54/3D7 parasite clone. We observed that the breadth of serological responses against 3D7 *var* /PfEMP1 domains affected the pattern of *var* expression at different time points in the CHMI. Exposed individuals with better ability to control infection in endemic regions had a broader breadth of response against 3D7 PfEMP1 domains, and had a distinct *var* expression pattern compared to individuals that were unsuccessful at controlling the infection. Conversely, in non-controller individuals, *var* expression amplified the breadth of response against 3D7 var/PfEMP1 domains, while controllers exhibit a comparatively stable response.

## 2 Results

### 2.1 Pre-existing immunity determines infection outcome

As previously described, 19 semi-immune adult males living in The Gambia were selected for their antibody levels against six *P. falciparum* antigens and classified as *sero-high* or *sero-low* [1]. All volunteers were inoculated with sporozoites (NF54/3D7 strain) by direct venous inoculation (DVI) and treated with anti-malarials once parasitaemia was detectable by microscopy or at the onset of symptoms. The infection outcome differed significantly in the two groups. *Sero-low* individuals had more symptoms (mean = 4.10, sd = 2.07) than *sero-high* individuals (mean = 5, std = 0.48) with symptoms occurring in both categories approximately 4-5 days after being positive for parasitaemia by PCR [1]. The peak parasitaemia before treatment in individuals classified as *sero-low* was significantly higher than the *sero-high* individuals (Mann Whitney U-Test; p = 0.01). In addition to the parasitaemia peak and clinical symptoms, we also compared the parasite growth rates across volunteers and the pre-patent period (Figure 1, Table 1), defined here as latency. Only two individuals had a long latency period (CH004 = 11 days, CH001 = 16 days), with one of them having an exponential rise in parasite growth rate post latency (CH004). The peak parasitaemia was lowest in the individual with highest latency period and lowest parasite growth rate (CH001). As in [3], we further used the differences in growth rate to classify the individuals as non-controllers (n=17) and controllers (n=2). The “Controllers” were defined as individuals with the longest latency and smallest parasite multiplication rate. Both individuals classified as controllers (CH001 and CH004) were *sero-high* (Figure 1 and Table 1).

**Figure 1:**
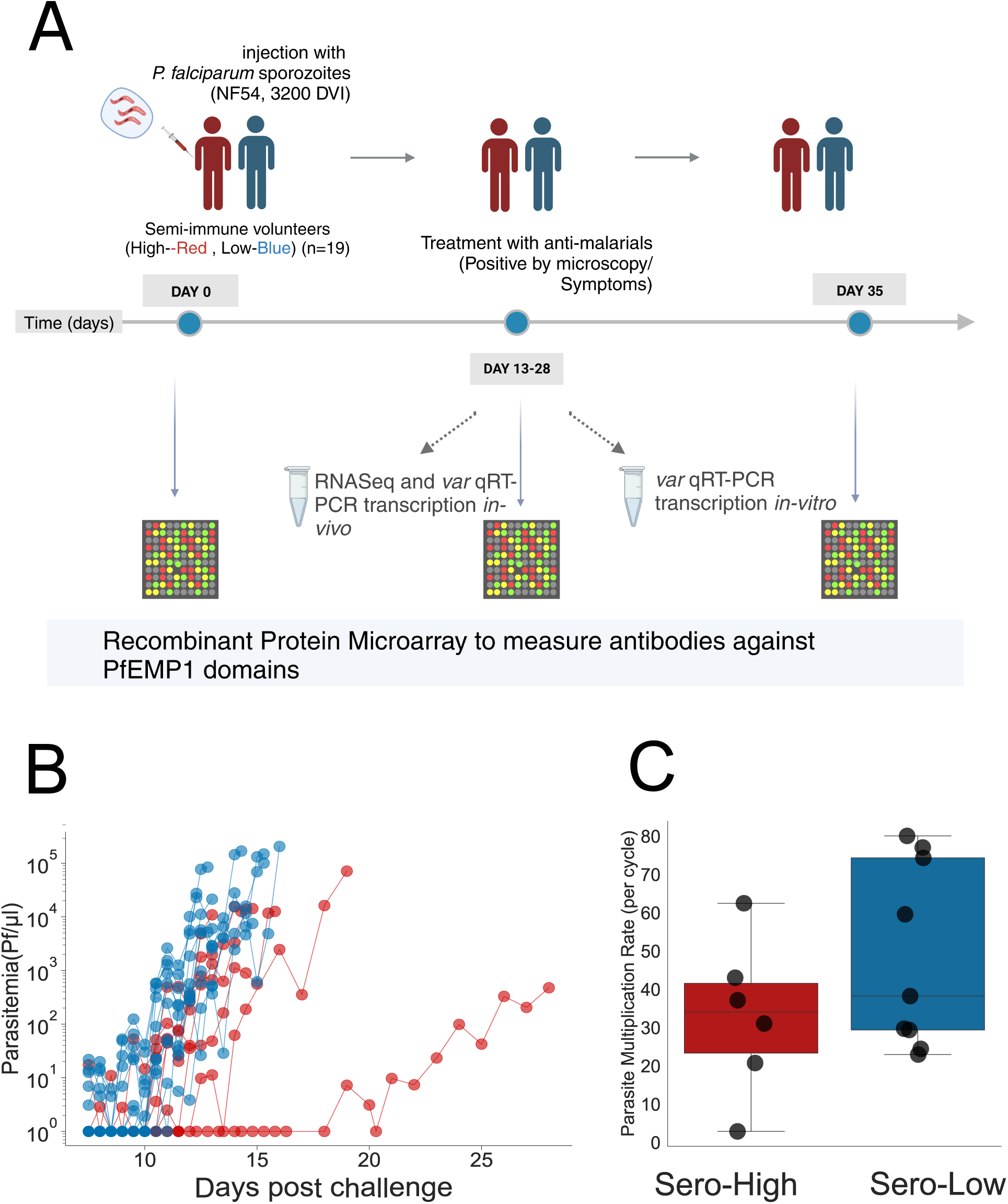
Characteristics of the CHMI for monitoring *var* expression and immune response: A) 19 semi-immune adult males were selected based on their pre-existing antibody levels against six *P. falciparum* antigens and classified as either *sero-high* (red) or *sero-low* (blue) [1]. All volunteers were infected with 3200 sporozoites (NF54/3D7 strain) on the same day and monitored for symptoms of malaria and parasitaemia, *var* gene expression and immune response against PfEMP1 protein domains. Venous blood samples were collected for *var* gene expression at one or two timepoints during the infection; on day 14 post inoculation and day of treatment/termination of study for “long” infections, or only on day of treatment in case an individual was treated before day 14 (short). The immune responses to PfEMP1 domains (n=158) were measured before, during (once or twice, depending on whether the individual was treated before or after day 14) and two weeks after the CHMI. B) parasitaemia vs days post infection, stratified as *sero-high* (red) or *sero-low* (blue). C) Parasite Multiplication Rates (fold change per cycle) across the groups of volunteers; classified here as *sero-high* (red) and *sero-low* (blue). DVI - direct venous inoculation.

**Table 1:** Parasitaemia characteristics for CHMI volunteers: Different parameters stratified by volunteer immune status before the start of the infection (*sero-low* or *sero-high*). The relationships between these parameters are highlighted in Figure 1. The latency period is defined as number of days until there is detectable parasitaemia. The generation time is the average time taken to double the parasite density. One sero-high individual (CH006) remained negative for the infection throughout the study, while another one (CH007) never reached a parasitaemia sufficient for transcriptome analysis.

| Volunteer | Immune Response before CHMI | Latency (days) | Entropy | Generation Time (days) | Peak parasitaemia (Pf/ml) |
| --- | --- | --- | --- | --- | --- |
| CH001 | sero-high | 16.00 | 1.95 | 1.30 | 476 |
| CH002 | sero-high | 10.45 | 3.00 | 0.38 | 1961 |
| CH003 | sero-high | 9.38 | 2.64 | 0.30 | 5187 |
| CH004 | sero-high | 11.37 | 1.60 | 0.46 | 10910 |
| CH008 | sero-high | 9.57 | 2.89 | 0.37 | 12645 |
| CH009 | sero-high | 9.48 | 3.58 | 0.34 | 14300 |
| CH010 | sero-high | 7.30 | 2.64 | 0.48 | 15400 |
| CH011 | sero-low | 9.38 | 3.30 | 0.32 | 21360 |
| CH012 | sero-low | 10.20 | 4.00 | 0.32 | 22880 |
| CH013 | sero-low | 6.39 | 3.85 | 0.41 | 27430 |
| CH014 | sero-low | 6.57 | 3.50 | 0.41 | 27980 |
| CH015 | sero-low | 6.81 | 3.40 | 0.43 | 71264 |
| CH016 | sero-low | 6.40 | 3.85 | 0.44 | 84475 |
| CH017 | sero-low | 9.21 | 4.06 | 0.34 | 100595 |
| CH018 | sero-low | 5.61 | 3.22 | 0.60 | 148950 |
| CH019 | sero-low | 8.94 | 3.69 | 0.38 | 168700 |
| CH020 | sero-low | 8.69 | 4.01 | 0.32 | 205850 |

### 2.2 VSAs are the main differentially expressed genes between sero-high and sero-low individuals

To understand how identical parasites adapt to different hosts, we performed a low input, whole transcriptome analysis of parasites recovered from ten volunteers on the day of treatment. Gene expression and subsequent differential expression analyses are highly influenced by the age and developmental stage of parasites [59]. Thus, we first estimated parasite ages from sequencing reads by calculating the maximum likelihood estimate against data from an *in vitro* time-course experiment [33]. Parasites were *∼* 7.6 hours post invasion (hpi) (95% CI; 6.67 – 9.35) (Figure S1 A & Supplementary file S1), with no significant disparity between the ages of *sero-low* and *sero-high* parasites (*p*=0.28; Student t-test) (Figure S1 B).

Applying a log2 fold change of > 2 (more than 4 fold difference) and Benjamini-Hochberg adjusted *p* value < 0.01 as cuttoff, 144 *P. falciparum* genes were differentially expressed, of which 103 were down-regulated and 41 up-regulated in *sero-high* compared to *sero-low*. Among these, genes coding for proteins with cell-cell adhesion predicted function were the most prominent, including 8 *var* genes (Figure 2 A, Supplementary file S2). Due to the nature of clonally variant gene transcription, a pooled analysis might not effectively capture VSAs expressed in a limited number of isolates. Consequently, we undertook pairwise comparisons of all isolates to detect genes displaying differential expression among individuals (Supplementary file S3). Genes were then ranked based on their frequency of appearance in these pairwise comparisons. Remarkably, the top 50 most differentially expressed genes exhibited a notable enrichment of *var* (8 out of the 61 *var* genes were present in the top 50) (Figure 2B). These observations indicated that the most variably expressed genes among individual volunteers were members of the *var* family. Other Variant Surface Antigens (VSAs) including *stevor* (*∼*30 members) and *rifin* (*∼*180 members), were poorly detected (Figure S2), likely due to their transcription peaking later in the intra-erythrocytic life cycle [31, 43, 68, 45]. The exception was the *rifin* PF3D7_0401600, which was detected in almost all samples (Supplementary Figure S2B, Supplementary file S4), and was also the major expressed *rifin* in a CHMI study with naive volunteers [38]. In contrast, *var* genes were robustly detected in all samples. To validate the accuracy of our *var* gene observations, we measured *var* expression in each volunteer by RT-qPCR and compared outcomes of both methods. A Bland-Altman plot comprising 610 observations (61 *var* genes in 10 volunteers) showed high c oncordance, with only 15 observations falling outside the 95% confidence interval (Figure 2C). Based on these observations, we focused our attention on *var* gene expression within hosts of varying immunity and the corresponding humoral immune responses.

**Figure 2:**
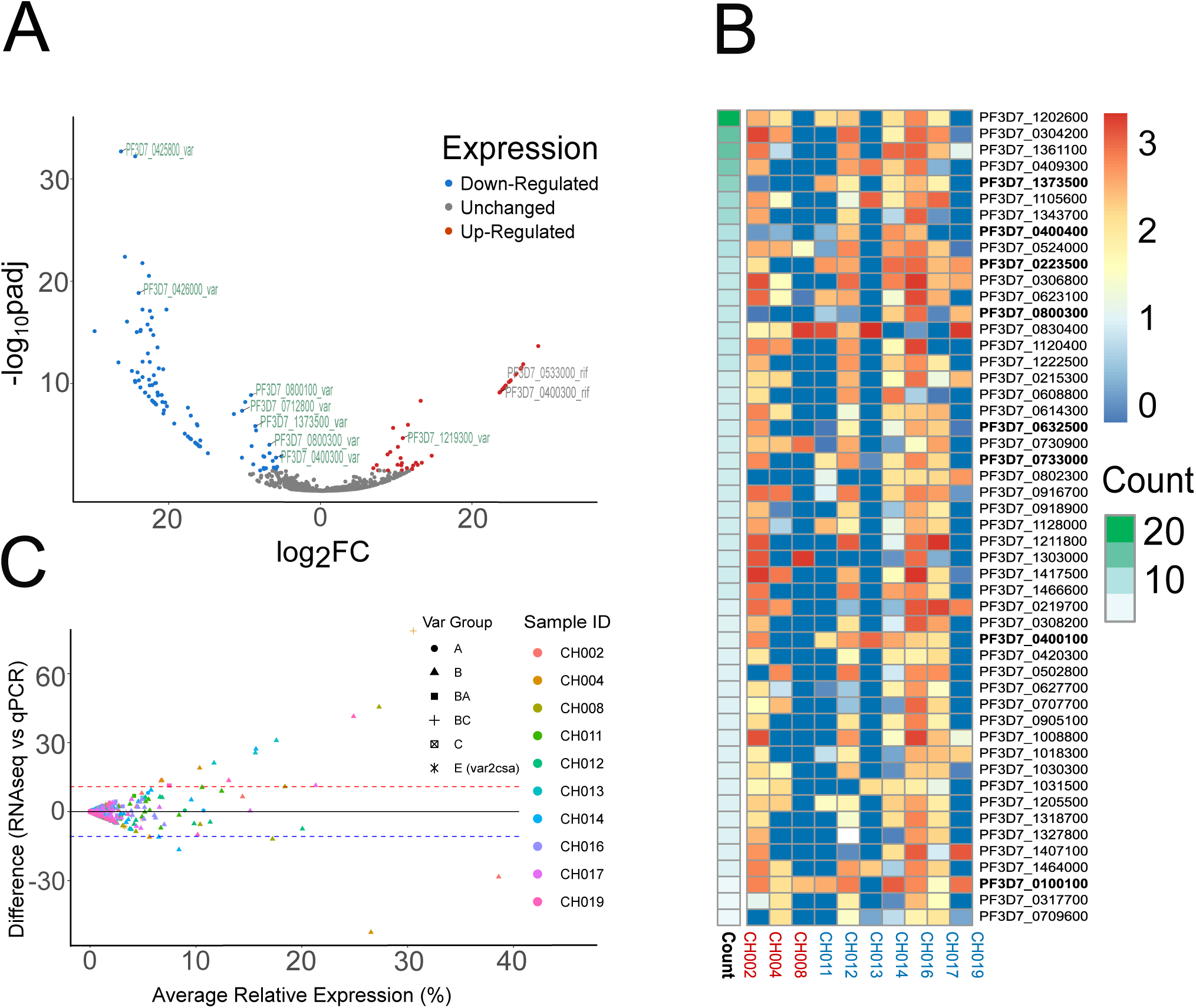
Whole transcriptome expression analysis: A) Differential expression analysis between sero-high and sero-low volunteers. Blue and red represent down and up-regulated genes, respectively. VSAs (*var* and *rif*) are labeled with gene IDs. B) Heat map of the log2 FPKM of the top 50 differentially expressed genes in a pairwise comparison. *Var* genes are in bold. Scale bar represent log2 fold change values. C) Bland-Altman plot of *var* expression by RT-qPCR and RNA sequencing.

### 2.3 *Var* gene expression pattern is shaped by host-immunity

To assess the impact of selection due to pre-existing immunity on *var* expression repertoire, *var* gene expression was analysed in all individuals by RT-qPCR. This was either done at day 14 post-inoculation and/or on the day of treatment. Comparison of total *var* quantity from RT-qPCR data showed higher *var* transcript levels in *sero-low* individuals (Mann-Whitney U-test, p-value <0.001). Hierarchically clustering of pooled volunteers from the current study and a previous one in Gabon [3], based on their *var* expression patterns alone showed distinct separation of controllers (CH001 & CH004 from The Gambian CHMI and L1.023, L1.26, L1.010, L.018, L1.028 from the CHMI in Gabon [3]) from *non-controllers* (Figure 3A).

**Figure 3:**
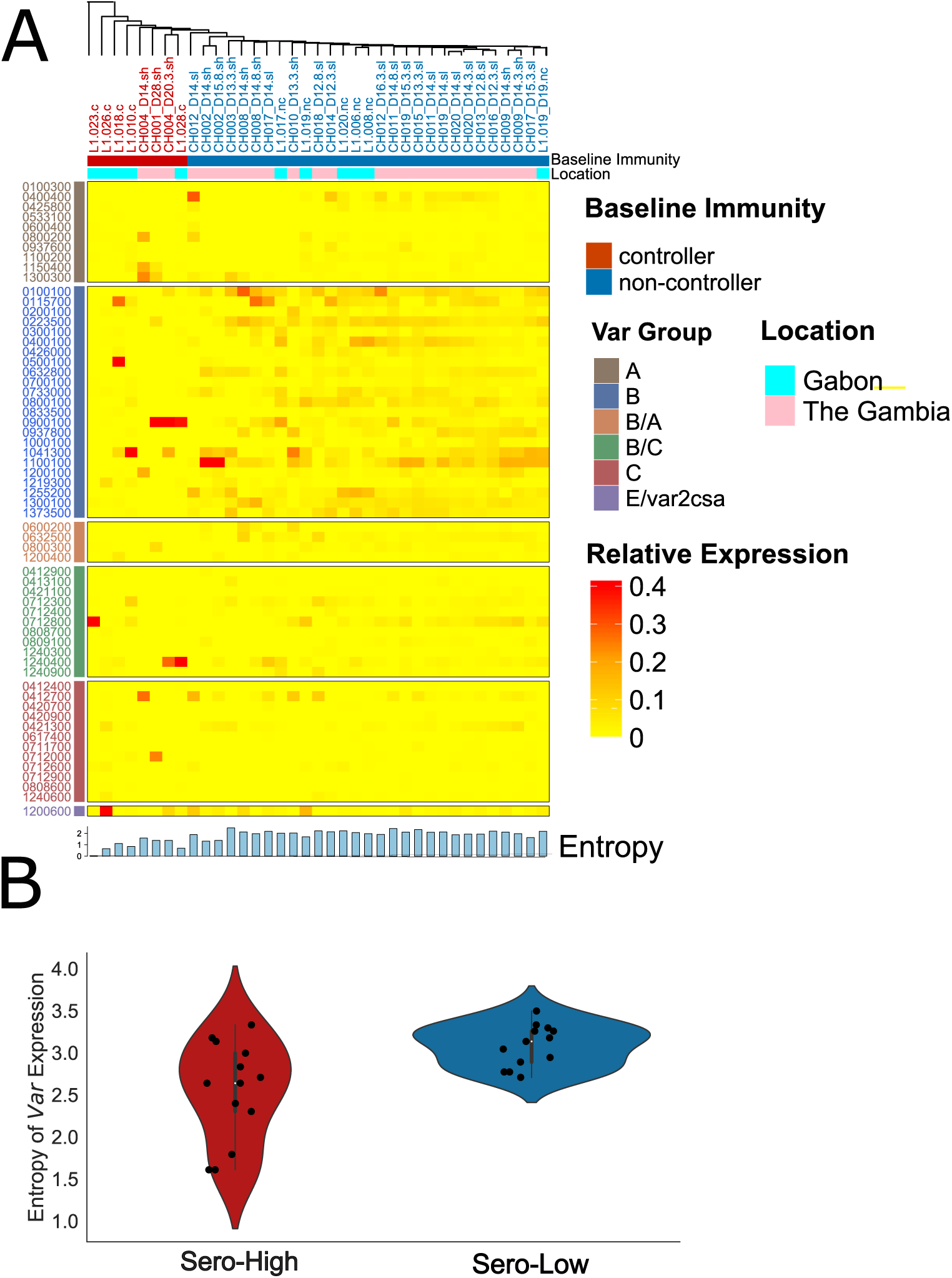
V*a*r expression landscape across two CHMIs. A heatmap of *var* gene relative expression proportion at several timepoints; includes data from this CHMI and from Gabon (at the day of peak infection). The relative expression of each gene is scaled with the total amount of *var* expression, quantified by RT-qPCR and ranges from low to high (0-0.40). The bottom of the heatmap is annotated by the entropy of expression, representing the amount and diversity of *var* genes in each sample. Top annotation is based on immunity and geographical region of origin. All volunteers across different timepoints were hierarchically clustered using an average-linkage method. Distinct clusters formed based on immunity, but not necessarily location. *Non-controllers* (blue) developed symptoms and patent parasitaemia faster than *controllers* (red) and expressed a high diversity of *var* genes during the CHMI, independent of the geographical region of origin of the volunteers. B) Entropy of *var* expression: violin plots for the Shannon index of *var* expression across the two categories of volunteers (*Sero-High*; red & *Sero-Low*; blue). In the Gambian cohort, the expression entropy was lowest for the two controllers: CH001 and CH004.

To estimate *var* gene heterogeneity, we compared the diversity of *var* repertoires across *sero-high* and *sero-low* groups by computing the Shannon entropy, a measure encompassing both diversity and relative abundance. Significantly higher entropies were observed for *sero-low* individuals (Mann-Whitney U-Statistic, p-value = 0.0002); indicating a larger breadth of *var* expression, in contrast to *sero-high* isolates that exhibited a more restricted repertoire of transcribed *var* genes (Figure 3B). This difference in diversity was also negatively associated with other markers of infection progression including latency (Spearmann Rank Correlation, r = -0.55, p = 0.02) and parasite doubling time (Spearmann Rank Correlation, r= -0.53, p =0.02).

In summary, the distinct (restricted) *var* gene expression pattern in *controllers* could be a proxy for a slow growing *P. falciparum* infection, possibly resulting from selection against specific PfEMP1s.

### 2.4 *In vivo var* gene renewal rates are very high

To evaluate *var* transcript profile changes *in vivo*, we compared the expression levels in individuals with two available timepoints, ranging from day 14 to day 20. We then computed the theoretical limits of probability to stop expressing a certain gene, given its expression at the first time point (the turnover probability of that gene). As expected, the turnover probability increased with time between day 14 post-infection and the day of peak infection (Figure 4 A). For the individual with the highest time gap between two time points (CH004), all genes expressed on day 14 were undetectable after three life cycles (6 days).

**Figure 4:**
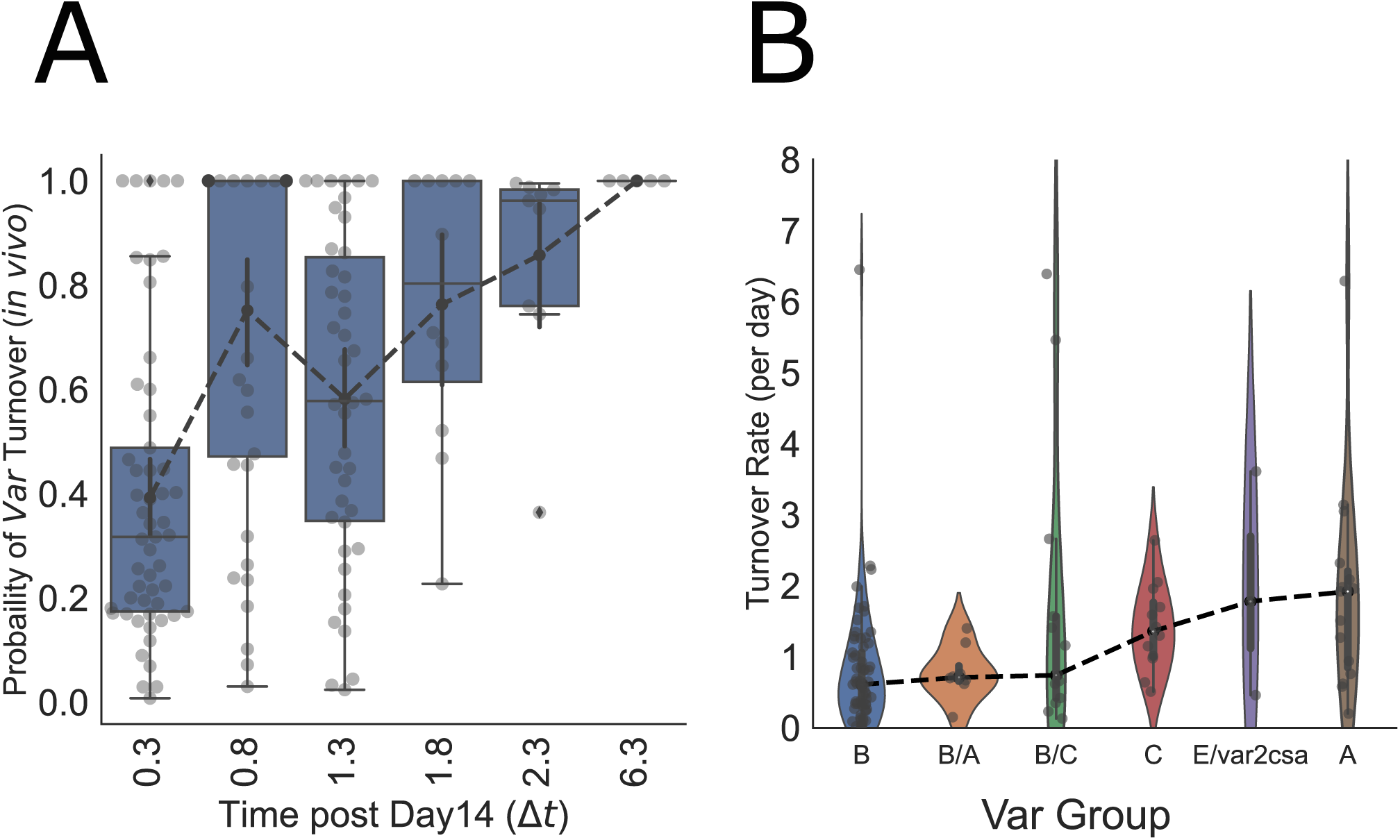
Quantifying change in *var* expression during an infection. A) Transition (turnover) probability comparison across different timepoints indicated that genes expressed at day 14 are no longer detectable by the end of the infection in most volunteers. This turnover probability is ordered between day 14 and day 20 post-infection. The median of the turnover probability is highest on day 20 (black dashed line). B) The finite-time transition (turnover) probability for each gene at each time point was used to calculate the instantaneous transition rate per day. The transition rates (off-rates) for various genes are shown per group, sorted by the median transition rate, in increasing order from left to right. Based on our data, Group A *var* genes had the higher transition rates *∼* 1.78 per day as compared to 0.59 per day for Group B *var* genes.

We further computed the instantaneous transition rates per unit time (days*^−^*^1^) during the CHMI, defined as turnover probability per unit time. On comparing the *var* transition rates per day across different *var* groups, groups A, C and E (*var2csa*) had the highest transition rates. (Figure 4 B). The median transition rates were higher than 1 per day in all three groups, which correspond to transition probabilities higher than 86% per cycle. These high *in vivo* transition rates may arise from a combination of the inherent switching rate and the potential selection pressure acting against parasites that express particular PfEMP1 variants.

### 2.5 *In vitro var* gene switching sustains a steady state with elevated entropy

Out of the original pool of volunteers (n=19), ten blood isolates drawn from seven volunteers were cultured *in vitro* for several cycles to investigate the impact of a lack of host-immunity on *var* expression patterns. Five isolates (CH001-D28, CH002-D14 CH002-D15.8 CH004-D14 & CH004-D20.3) belonged to the *sero-high*, while five isolates (CH012-D14, CH012-D16.3, CH014-D12.3, CH016-D12.3 & CH020-D14)^1^ were from the *sero-low* category. For each isolate in culture, we analysed *var* transcription profiles by RT-qPCR at 3 to 8 time points for up to 100 days (Figure S3). Isolates derived from sero-low individuals all converged to near-identical *var* gene expression pattern within 20 days in culture, with all Pearson correlation values above 0.80 (Figure S4). In contrast, sero-high isolates did not demonstrate any uniform expression pattern even after 50 days of culture, with Pearson values ranging from 0.20 to 0.95.

To rigorously assess the hypothesis that the host immunity level may have a lasting impact on *var* gene expression *in vitro*, we integrated data analysis with mathematical modeling.

First, the multi-dimensional traces from the ten timeseries were projected onto a 3D space using PCA. We divided the time points in three categories: early (0-6 days), intermediate (20 *≤* 50 days), and late (50-100 days). The projected point clouds from different categories show that the *var* gene global dynamics is quantitatively different for samples from *sero-low* and *sero-high* volunteers (Figure 5). For samples derived from *sero-low* volunteers, *var* gene transcription profiles are more spread at early time points than late time points (Figure 5A), shown quantitatively by using the convex hull of the PCA projected points (Figure 5C). This suggests that after a fast transient phase, the *var* gene transcription profile reaches a steady state; an observation which is similar for all isolates. This can also be seen in Figure 5E where the transcription profiles of all late timepoint samples are compared without projection. *Sero-high* volunteer samples showed a different trend: the variability of the transcription programs starts from smaller values, but not all samples reached a steady state (the final variability is not close to zero) (Figure 5B,D,F). The similarity of the late time *var* gene expression profiles in different samples was also tested to be higher for *Sero-low* compared to *Sero-high* volunteer samples by using an AUC criterion (Figure S5). Nevertheless, when utilizing entropy as a metric to assess the diversity of *var* gene expression within a particular profile, we observed that all samples exhibited a consistent trend of monotonic entropy increase with the highest entropy being reached in the steady state (Figure 5I). Moreover, the primary distinction between *sero-low* and *sero-high* samples appears to be the initial transcription profile, which exhibits higher entropy in the former as opposed to the latter.

**Figure 5:**
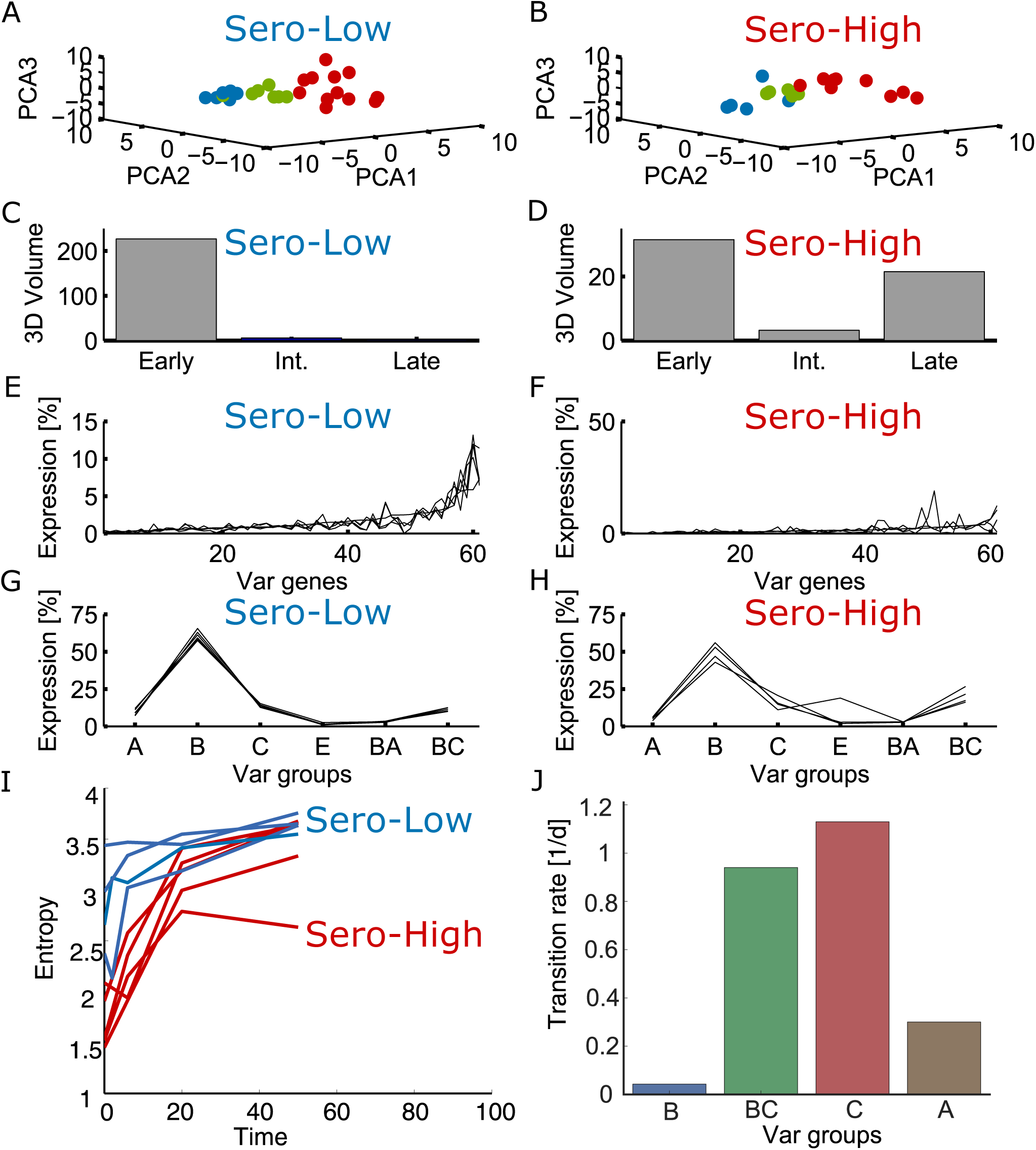
In vitro dynamics of *var* gene transcription. A,B) distribution of early (red), intermediate (green) and late (blue) time samples projected on the 3 first PCA components. C,D) volume of the convex hull of the samples projected on the same 3D space was used as a proxy for the variability of the *var* transcription programs. E,F) probabilities of expression of different *var* genes for all late time samples. G,H) probabilities of expression of different groups of *var* genes for all late time samples. I) *in vitro* evolution of *var* gene expression entropy. J) estimated *in vitro* transition rates.

Secondly, we performed a more accurate analysis of the *var* gene transcription dynamics using a mathematical model. In the absence of host immunity, the *in vitro* dynamics is not influenced by selection, but results only from intrinsic switching of the *var* gene system. Under this hypothesis, the *var* gene dynamics can be modelled as a continuous time, four-state Markov chain (see Methods and Materials). The four-states Markov chain model fits well to the data of all the samples (Figure S7). As observed in the first analysis of the data, the theoretical steady state of the inferred Markov chain has high entropy, where many *var* genes are significantly expressed. *Sero-high* samples require more time to reach steady state due to distinct initial data compared to the steady state, while *sero-low* samples have initial states with higher entropy closer to the steady state. The fitted values of the transition rates are represented in the Figure S6. The *C* and *BC* switching rates are similar to turnover transition rates *in vivo*, while those for *A* and *B* are much smaller than the respective *in vivo* rates (Figure 5J). Altogether, our data indicate that immune selection within sero-high individuals may still impact the *var* gene transcription profile *in vitro* after 25 generations.

### 2.6 Immune responses against VSAs differ significantly between sero-high and sero-low individuals

We evaluated the antibody levels against different antigenic domains corresponding to PfEMP1s and other antigens by exploiting recombinant protein microarray data. Serological responses to a total of 213 *falciparum* PfEMP1 domains (159 from 3D7, 54 from three other strains) and 18 other recombinant antigenic proteins (MSP1, CSP, AMA1, RIFIN and STEVOR) were analysed for each volunteer at several timepoints: before the injection (day 0), during the infection (Range: day 13-28) and about 2-3 weeks after treatment (day 35) (Figure 6). At day 0, antibodies against non-PfEMP1 surface antigens were associated with the latency period (AMA1: r= 0.80, *p <* 0.01 & MSP1: r = 0.48, *p <* 0.01 and the peak parasitaemia (AMA1: r= -0.80, *p <* 0.01, MSP1: r= -0.59,*p <* 0.01) using Spearmann Rank Correlation. These results confirmed previous data from the same CHMI study [1].

**Figure 6:**
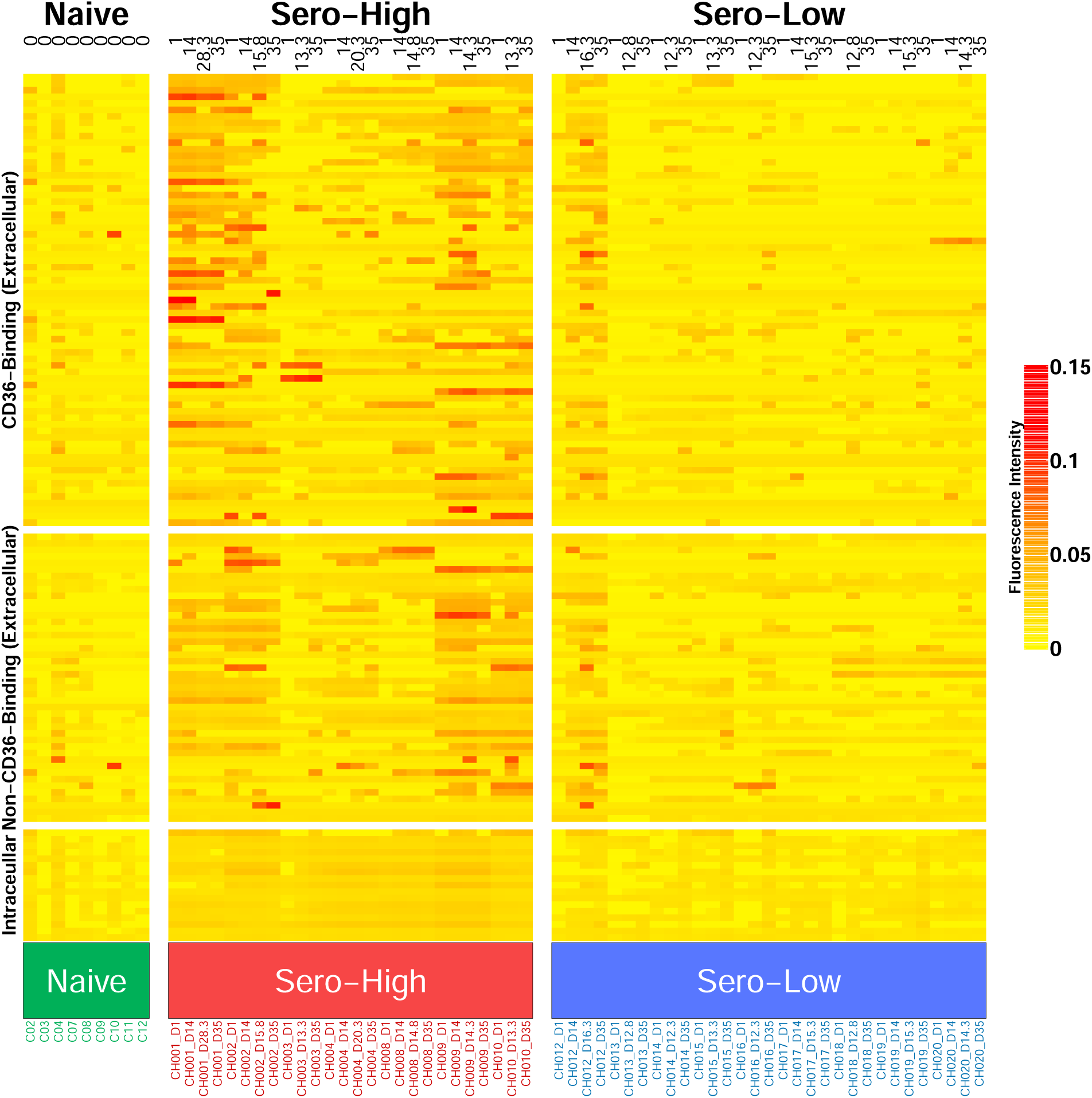
Heatmap of serorecognition of all PfEMP1 domains from all volunteers. Each column is a time point for a particular volunteer during the infection (top annotation). The *sero-high* and *sero-low* individuals have been annotated in green and blue, respectively. The naive individuals are grouped on the left (labelled and annotated in green). Each row corresponds to a PfEMP1 domain, grouped based on the receptors they bind to and the fluorescence intensity is scaled for each domain, in increasing order (from yellow to red). The domains are grouped by the type of antibody binding types they are associated with, namely; CD36 Binding domains (associated with less severe symptoms), Non-CD36 Binding domains (EPCR-binding) and Intracellular domains (ATS)

Moreover, based on the breadth of response at the beginning of the CHMI, *sero-high* individuals had sero-recognition of a significantly higher number of 3D7 PfEMP1 domains in comparison to *sero-low* individuals before (Mann-Whitney U-Test, *p <* 0.01) and after the infection (Mann-Whitney U-Test, *p <* 0.01) (Figure 7 A). When examining the change in the number of recognized domains before and after the CHMI study, no significant induced response was observed in the *sero-high* group (Mann-Whitney U-Test, *p <* 0.01) (Figure 7A). This is in sharp contrast with the breadth of antibody levels in sero-low individuals, that more than doubled after the infection (mean number of recognized domains pre-infection *∼*11, *±*6; post-infection *∼*28, *±*12, Mann-Whitney U-Test= 17.0, *p <* 0.01).

**Figure 7:**
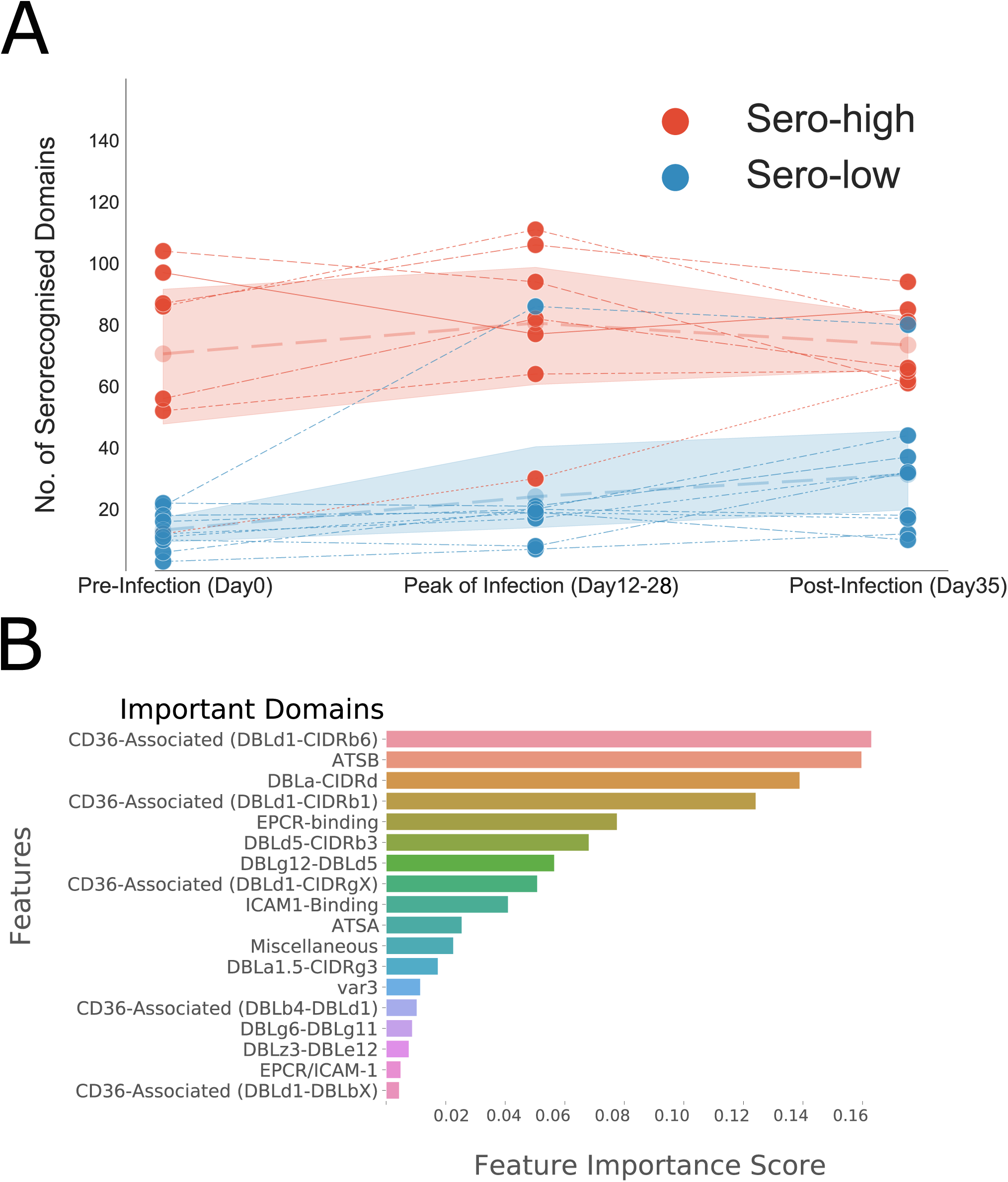
Significant increase of serorecognised PfEMP1 domains post-CHMI in *sero-low* individuals. A) Sero-positivity throughout time in CHMI participants (top panel): The scatterplot showing the number of domains that were sero-recognized at different timepoints during the CHMI across the two groups, *sero-high* (red) and *sero-low* (blue). The shaded region represents the confidence interval (*±* 2 SD) for the number of domains recognized in each sub-group. B) Relative feature importance score to predict infection outcome. The random Forest classification model was ranked according to the association between antibody levels against domain groups to predict volunteer subgroups.

Additionally, we employed a random forest model to predict the subgroup to which the volunteers were linked using microarray data related to PfEMP1 domain responses. Using this model, the subgroups were re-confirmed with a predictive accuracy of 98% (Figure 7B). Feature scores of the model revealed that the most important domains for predicting infection outcome were either DBL-CIDR di-domains, ATS domains or CIDR*α*-binding domains, with CIDR*α*-binding domains being the sub-domains that are linked to severe symptoms of *falciparum* infection. The sero-recognition breadth within each individual was negatively correlated with infection markers *in vivo*, i.e. latency (Spearmann Correlation: 0.67,*p <* 0.01) and the peak parasitaemia (Spearmann Correlation: -0.68,*p <* 0.01) (Figure S8).

### 2.7 *Var* expression is moderately associated with pre-existing specific immune response

To investigate the interplay between pre-existing specific immune responses and PfEMP1 expression, we examined our primary hypothesis that the presence of antibodies targeting specific PfEMP1 domains would impose a selective pressure against infected red blood cells (iRBCs) expressing the corresponding *var* gene during the asexual blood stage. Overall, we observed a moderate association between the breadth of PfEMP1 domain recognition prior to challenge and the Shannon entropy of *var* gene expression during CHMI (day 12-20) (Spearman Rank Correlation = -0.52, *p <* 0.05). This suggests that the diversity of *var* gene expression may be constrained by the breadth of immune responses directed against PfEMP1 domains.

To make inter-PfEMP1 responses throughout the CHMI comparable, we used a discretized method to evaluate both the fluoroscence intensity values for antibody levels per domain and the expression levels of the *var* transcripts (described in Methods and Materials) based on quantiles per domain (for immune response data) and per gene (expression data). On comparing the intensity of recognition at the start of infection to *var* gene expression, we observed a moderate association during the CHMI (Figure 8). This relationship was strongest for *var* groups ’B\C’,’A’ and ’B’, indicating that some PfEMP1 expression could have been inhibited by existing specific immune response. However the antibody intensity against PfEMP1s corresponding to the intermediate group ’B/A’ and ’var2csa’ was not significantly associated with reduction in expression during the CHMI.

**Figure 8:**
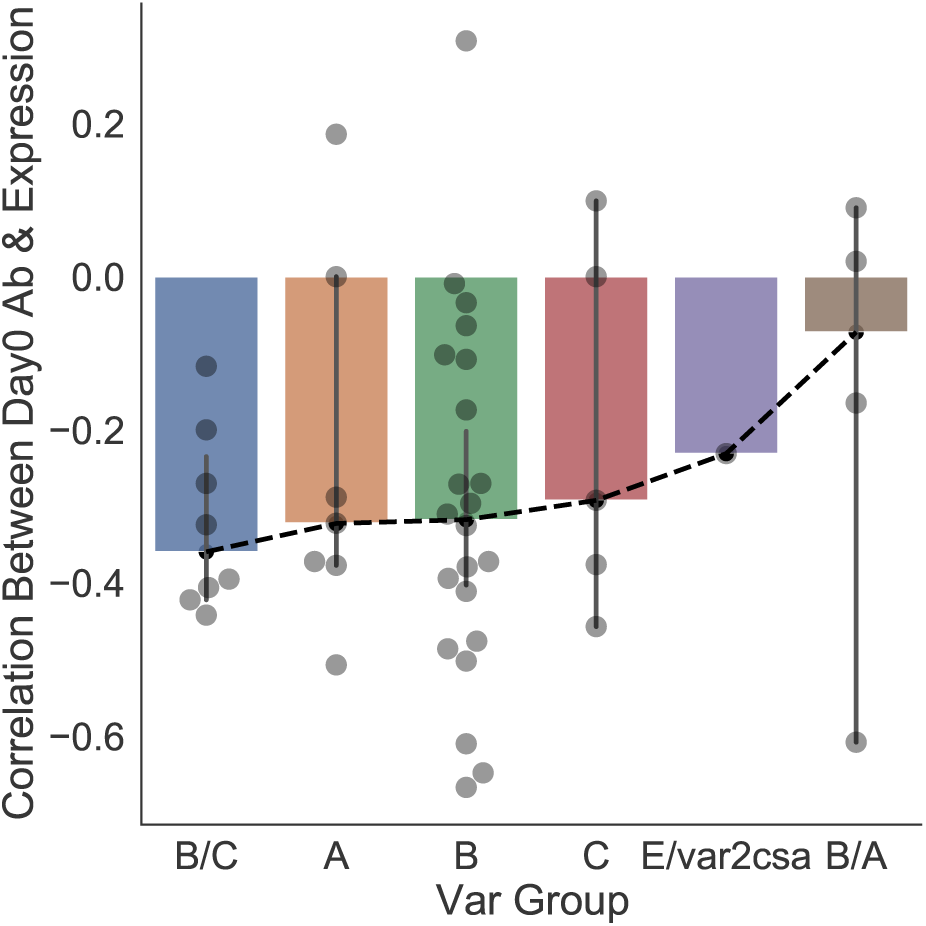
Anti-correlation between pre-existing PfEMP1 antibodies on day 0 and *var* expression during the CHMI. A PfEMP1 was considered sero-recognised in an individual if at least one of its domains showed antibody signal on the array. We calculated, across all volunteers, the Spearmann Rank correlation between the PfEMP1 sero-recognition intensity on day 0 and its subsequent expression during the infection. The correlation was plotted separately for each *var* group, ordered by the median of the correlation between the two quantities.

### 2.8 The increase in antibody levels is not necessarily dependent on the intensity of PfEMP1 expression during the CHMI

In order to examine the hypothesis that the overall intensity of PfEMP1, specifically the quantity of PfEMP1 expressed at a particular time point within an individual, initiates antibody production, we contrasted the discrete intensity levels at which various *var* genes were expressed during CHMI with the quantity of antibodies acquired against the corresponding PfEMP1. On evaluating the correlation between these two variables, we observed no evident of association in antibody development towards genes expressed at high intensity when compared group-wise for all *var* groups (Figure 9). These results indicate that the gain in antibody levels is not necessarily dependent on the intensity with which a particular gene was expressed during the infection.

**Figure 9:**
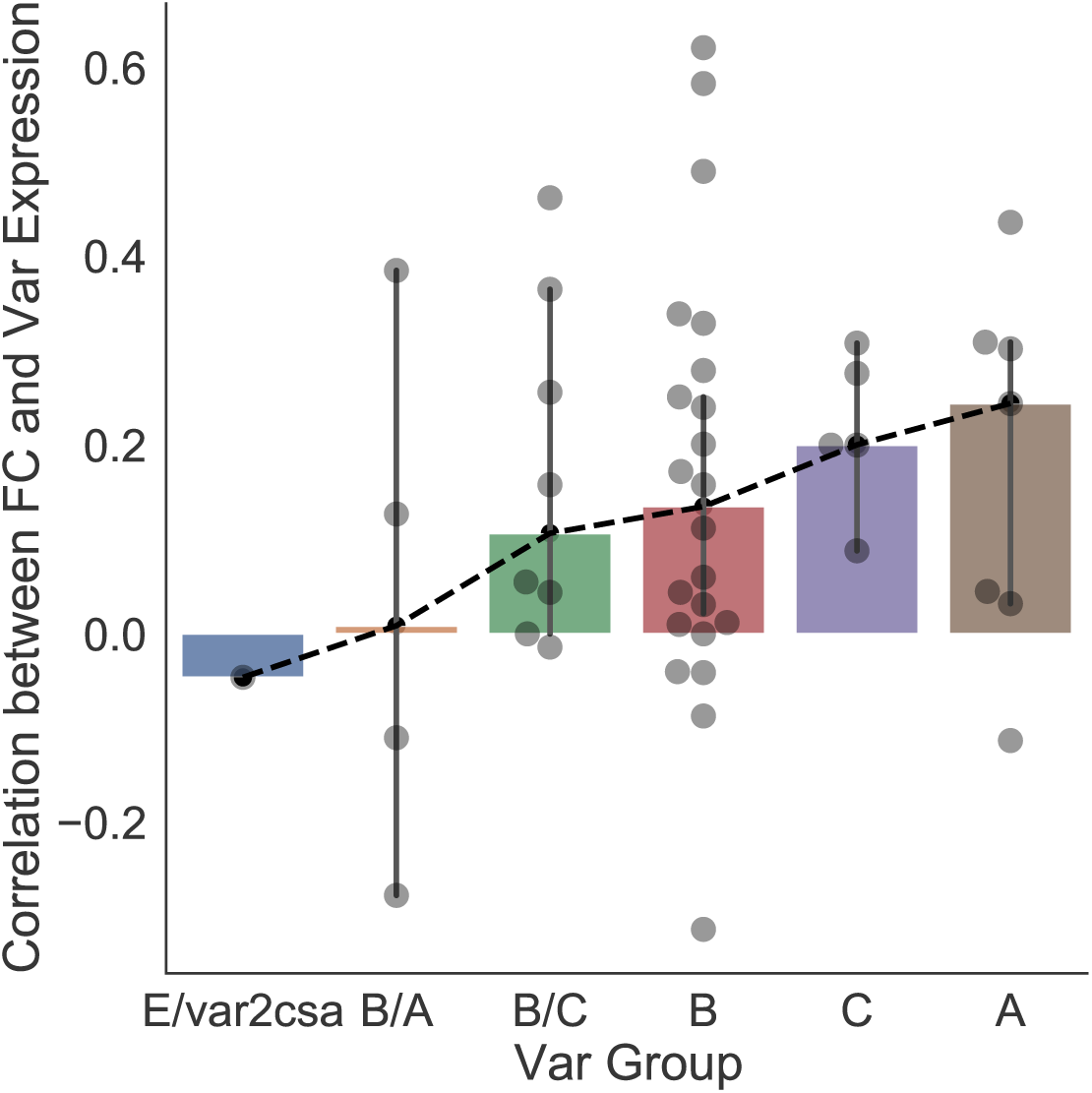
Correlation between expression intensity of PfEMP1s and antibody acquisition at day 35. Group-wise comparison of *var* genes based on the correlation between their expression intensity during the infection and the corresponding antibody gain against them at day 35.

## 3 Discussion

Controlled Human Malaria Infection (CHMI) studies are useful not only for evaluating drug efficacy and vaccine development but also for understanding pathogenesis of *Plasmodium*within the human host and its interactions with the immune system in real time [55]. Leveraging clinical and experimental data from CHMI studies, our aim was to document the heterogeneity in host responses and compare infection characteristics that transcend different geographical regions.

Individuals selected for their previous malarial exposure showed diverse infection outcome and *var* expression heterogeneity, consistent with previous findings in Gabon [3]. In our study, the whole transcriptome sequencing approach underscores *var* genes as the primary cause of transcriptome heterogeneity among isolates. Despite the geographical distance between Gabon and The Gambia, there is a consistent *var* gene expression pattern linked to pre-existing immunity against malaria. The observed similarity suggested an immune response that selectively targeted certain PfEMP1 variants in individuals classified as “controllers”. These individuals, characterized by low entropy profiles featuring limited expressed genes, exhibited longer latency periods, lower parasitaemia levels before treatment, and demonstrated recognition of a wide array of PfEMP1 domains prior to the infection challenge. On the other hand, individuals classified as “non-controllers”, who had high entropy profiles and a wide range of expressed genes, showed elevated parasitaemia levels, shorter infection delays, and limited recognition of PfEMP1 domains. This is reminiscent of the rodent malaria model, in which *P. chabaudi* parasites express a very limited number of *pir* genes in chronic infections [10].

Antigenic variation, encompassing intrinsic switching of the PfEMP1 proteins that a parasite displays at the red blood cell surface, is regarded as a prominent survival strategy employed by *P. falciparum* during its blood cycle. This process has been extensively studied *in vitro* [27, 48, 70]. Nevertheless, antigenic variation is also contingent on the host-pathogen interaction, triggered both by intrinsic switching and by elimination of variants through the immune system’s negative selection process. Negative-selection eliminates variants and is therefore detrimental to the parasite survival, whereas intrinsic switching replaces one variant with another and maintains the expression diversity of the antigens. Here, from 9 volunteers, two blood isolate timepoints were available, providing a rare opportunity to quantify the turnover rates of *var* genes *in vivo*. Turnover probabilities rapidly increased over time. For instance, in the volunteer that was able to control the infection until the end of the CHMI without turning symptomatic, we found that the initial repertoire of *var* genes had completely changed after three cycles (between day 14 and 20). Given the short interval between timepoints, this shows that the *var* turnover during an infection is much higher than reported previously [42, 20]. Our mathematical estimates based on a Markov chain model lead to turnover probability estimates with group averages between 69% and 97% per generation (corresponding to instantaneous rates from 0.59 to 1.78 per day, see Figure 4B), likely as a result of a combination of intrinsic switching and immune selection. Additionally, group A *var* had the highest rates of transition *in vivo*, a phenomena previously observed *in vitro* [43].

To get an accurate estimate of switching events and rates in the absence of immunity, we modelled the *var* transcripts coming from the CHMI volunteers *in vitro* for several cycles in culture. In our study setting, although smaller than *in vivo* values, intrinsic switching rates were found higher than reported elsewhere [51, 27]. *In vitro* transition (switching) probabilities for *var* genes B and A (7% and 44% per generation, respectively) are over two-fold lower than *in vivo* turnover, whereas the *in vitro* values for var C and BC (57% and 49% per generation, respectively) are close to *in vivo* values. These large, per group values, do not exclude that some rarely transcribed *var* genes have low transition rates, even within fast groups. Furthermore, *var* expression profiles evolved *in vitro* towards a steady-state distribution marked by a significant increase in entropy and many expressed *var* genes. The steady state distribution in both sub-groups comprised as major constituents *var* groups B and C, that is compatible with previous findings 43. In the sero-low groups, the parasite populations reached the corresponding steady state relatively quickly, within fewer than 10 cycles. Remarkably, for sero-high individuals, the parasite cultures took longer to converge to the steady state, with some samples retaining memory of the initial state even after 50 days in culture. We therefore hypothesize that epigenetic imprinting on parasite populations from hosts with robust immune responses led to a *var* gene repertoire significantly divergent from the *in vitro* steady state. However, given that the switching rates were found to be similar, the disparities in time to reach a steady state can be primarily attributed to differences in the expression profiles between the two groups at the peak of infection.

We propose that in all sero-low individuals expressing multiple *var* genes, the *in vivo var* distribution closely mirrors the *in vitro* steady state. This is consistent with the hypothesis that *var* gene expression is reset during mosquito and/or hepatic stage passage [4]. We also hypothesize that a similar state is initially present but is rapidly lost through negative selection in sero-high individuals. This state is reached by parasite intrinsic *var* gene switching as part of a “bet-hedging” strategy in a hostile environment. In all individuals, the observed distribution at later *in vivo* time points evolve to a less diverse repertoire through negative selection. Several studies highlight that the initial probabilities of expression of different variants at the onset of blood stage infection are consistent across multiple CHMI studies with limited stochasticity between individuals [4, 26]. : Similarly, broad group B *var* expression was observed in a CHMI study with the 7G8 clone, although in this instance, a group C *var* was the most dominantly expressed in all volunteers [69][45, 69].

The most common approach to study naturally acquired immunity involves incubating antibodies from semi-immune individuals against parasite recombinant proteins derived from the 3D7 strain [12, 46]. However, to test antibody sero-recognition against the PfEMP1 family, these assays must rely on some level of cross-reactivity, due to the extremely high *var* gene sequence polymorphism. Here, we circumvented this limitation with a 3D7-based protein array and plasma samples from 3D7-infected individuals, allowing us to determine the precise antibody-acquisition against each PfEMP1 variant. The sero-low/sero-high grouping made from Luminex assay [1] was validated with MSP1, CSP and AMA1 on our protein array. As expected, these *sero-high*individuals also sero-recognised a significantly larger proportion of PfEMP1 domains. These pre-existing antibodies decreased the likelihood of reaching the treatment threshold for the infection. The diversity in recognition, coupled with cross-reactivity against VSAs can be considered crucial to prevent symptomatic infections [11, 30, 24] for controlling malaria infections. Our results re-confirm that anti-PfEMP1 immunity is a marker for infection outcome and severity of malaria infection [66, 8], and that certain PfEMP1 subsets have been linked to shield against severe symptoms of the disease [58, 60, 2, 41]. We also found that the pre-existing specific PfEMP1 antibody levels were at least moderately negatively associated with the expression of *var* genes during the infection, and this stems from acquired immunity to previous malarial infections. The negative correlation was most pronounced for group B and A PfEMP1, which constitute the *var* gene repertoire in early stages of infection.

Although blood-stage infections were artificially shortened by anti-malarial drugs, the breadth of antibody levels against PfEMP1 domains drastically increased by day 35 in sero-low individuals. Among the notable increases in antibodies was against the PfEMP1 domain ATS, located inside the red blood cell. This finding has been consistently demonstrated [61, 46], establishing ATS, the only conserved PfEMP1 domain, as a marker of exposure rather than protection. We did not identify a direct correlation between the *var* genes detected during the infection and the antibody-acquisition by day 35. Merozoites emerge from the liver around day 6, thus the *var* gene expression over the first 3 cycles is unknown, but they could have led to acquisition of novel antibodies observed post-infection. The observation that many highly expressed *var* genes at peak parasitaemia did not trigger an antibody response is intriguing. We hypothesize that certain PfEMP1 domains are poorly immunogenic. While it is evident that PfEMP1s generally induce a robust immune response, in fact most antibodies targeting surface antigens are against PfEMP1 [12]. However, whether surface expression of a PfEMP1 automatically generates a new antibody had not been previously explored. We argue that *P. falciparum* has evolved not only an extremely polymorphic gene family but also protein domains that are relatively weakly immunogenic. This evolutionary pressure does not apply to the intracellular ATS domain, explaining its higher immunogenicity.

In summary, our findings corroborate the following scenario: The driving force behind maintaining a high-entropy repertoire of PfEMP1 variants is the intrinsic *var* gene switching. The establishment of a high-entropy repertoire occurs through resetting during mosquito and liver passage. This bet-hedging strategy proves to be effective when the parasites are confronted with a less diverse immune response. The high entropy repertoire is maintained by intrinsic switching in *sero-low* individuals, but does not survive negative-selection in *sero-high* individuals. However, some PfEMP1 domains are only poorly immunogenic and can persist. As such, poor immunogenicity combined with bet-hedging insures the parasite survival during CHMI. Intrinsic *var* gene switching is responsible for the reset and maintenance of a diverse *var* repertoire. Its high rates represent a challenge for the immune system during CHMI, because of the limited immunogenicity and duration of the infection. In the case of prolonged infections, such rapid rates of exhaustion of the repertoire could be a disadvantage for the parasite, which then has to depend on alternative mechanisms to generate variability such as recombination [64, 16, 15]. Interestingly, parasites submitted to high negative selection in *sero-high* individuals tend to express more stable variants and recover much slower, the high entropy steady state. This again could work in favor of the host/immune system and contribute to establishing a dynamic asymptomatic equilibrium during extended infections.

## 4 Methods and Materials

### 4.1 Epidemiological study design and sample collection

The epidemiological data used in this study was obtained from a previously published, non-randomized clinical trial in the Gambia (low transmission intensity) carried out by the Medical Research Council Unit The Gambia (MRCG) [1]. Briefly, participants aged between 18-35 years were recruited for this study and were screened for various hematological and biochemical abnormalities. Previous malaria exposure in these participants before DVI (Direct Venous Inoculation) was approximated using the LUMINEX platform by comparing responses against 6 malaria antigens (AMA-1, MSP1.19, GLURP.R2, GEXP18, Etramp5, Rh2) known to be markers of malaria exposure. Based on these responses, the volunteers were classified into two groups, *sero-high* and *sero-low* [1]. All volunteers received PfSPZ Challenge (3.2 × 103 PfSPZ in 0.5 mL, NF54/3D7 strain from Sanaria) by direct venous inoculation. Venous blood samples were collected the day before the inoculation (Day 0), once or twice between day 11 to 28, and at at day 35. All volunteers were treated with artemether-lumefantrine once thick blood smears were positive with *P. falciparum*.

### 4.2 Parasite enrichment and sorting

Infected venous erythrocytes frozen in glycerolyte were thawed and immediately treated with Streptolysin O (SLO) from Streptococcus pyogens (Sigma) as previously described [28, 9], with few modifications. Briefly, lyophilized SLO was reconstituted at 25U/µL stock concentrations, activated at room temperature for 15 minutes with 1M dithiothreitol (DTT) and used at a final activity of 1U/µL. Cells were lysed at room temperature for 6 minutes and reaction quenched with 5% PBS-BSA. Cell pellets were resolved on 60% Percoll gradient to remove cell debris, by centrifugation at 2500 x g for 3 minutes. Pellets were washed twice with PBS and stained with 500µL of 1:2000 dilutions of Vybrant DyeCycle Green Stain (Thermo Fisher) for 30 minutes at 37oC. Where possible, 100 infected erythrocytes were sorted in triplicates with a BD FACSAria flow cytometer (BD Biosciences) into wells containing lysis buffer of 2µL 0.8% Triton-X100, 1µL of 10mM dNTP mix (Thermo Fisher), 0.1µL of 20U/µL RNase Inhibitor (SUPERase•In™; Thermo Fisher), 0.1µL of 100µM non-anchored oligo dT (IDT) [50], 0.4µL of 50% polyethylene glycol (PEG8000) (Sigma) and 0.4µL nuclease-free water. Plates were snap-frozen on dry ice and stored at -80oC until use.

### 4.3 Whole transcriptome amplification with SMARTseq2

Complementary DNA (cDNA) were synthesized from sorted cells with a modified version of the SMARTseq2 protocol which has been optimized for *Plasmodium* [50], with few modifications. Briefly, a molecular crowding step [5] was included to improve library yield by adding 5% polyethylene glycol (PEG8000) (Sigma) to the lysis buffer [25]. Additionally, the SmartScribe (Clontech) reverse transcriptase was substituted with the better performing Maxima H (Thermo Fisher) at the cDNA synthesis step [5, 25, 72]. Frozen plates were thawed and incubated at 72oC for 5 minutes. cDNA synthesis master mix with final concentrations of 1X Maxima H RT buffer, 10µM TSO (Qiagen), 5U SUPERase•In RNase Inhibitor, 25U Maxima H enzyme, and nuclease-free water in 6µL volumes were added to each well. The cDNA was synthesized at 42oC for 90 minutes, followed by 10 cycles of 2 minutes at 50oC and 2 minutes at 42oC, and deactivation at 85oC for 5 minutes. cDNA was then amplified at 26 cycles with the KAPA HiFi HotStart ReadyMix PCR Kit (KAPA Biosystems), using the following conditions: denaturation at 98oC for 3 minutes, cycling steps of denaturing at 98oC for 20 seconds, annealing at 67oC for 15 seconds, extension at 72oC for 6 minutes, and final extension at 72oC for 5 minutes. PCR products were cleaned with 1X AMPure XP beads (Beckman Coulter), and eluted with 20µL of nuclease-free water. cDNA quantity and quality were assessed with Qubit dsDNA HS Assay (Invitrogen) and Agilent High Sensitivity DNA Assay (Agilent), on a Qubit 4 fluorometer and Agilent 2100 Bioanalyzer, respectively.

### 4.4 Whole transcriptome sequencing and data analysis

Amplified whole transcriptomes were sequenced by BGI genomics (Hong Kong). Paired-end fastQ files were aligned with HISAT2 (default alignment parameters) [29] and bam files made with SAMtools [34]. The SummarizeOverlaps feature of the GenomicAlignments package [32] was used to count reads against the *P. falciparum* 3D7 reference genome (version 3.0) and DESeq2 [36] used for differential expression analysis in R.

### 4.5 Parasite culturing

Cryopreserved parasites were thawed with NaCl solution [56], and parasites cultured in RPMI-1640 (sigma) supplemented with 25mM HEPES, 2mM L-glutamine, 0.5% Albumax II (sigma) and 50µg/L gentamicin (sigma). Parasites were cultured in 10mL volumes at 2% haematocrit in a blood gas environment of 90% N2, 5% CO2 and 5% oxygen. Parasites were harvested for RNA extraction at respective timepoints after synchronization with 5% D-sorbitol.

### 4.6 RNA extraction from samples stored in RNAprotect and cultured isolates

Total RNA was extracted by the phenol-chloroform extraction method with TRIzol reagent. For *in vivo* samples stored in RNAprotect Cell Reagent (Qiagen), five volumes of TRIzol reagent (Ambion) was added for homogenisation. For *in vitro* samples, 1mL of TRIzol was added to 200µL erythrocyte pellets and homogenised. One-fifth TRIzol volumes of chloroform (Sigma-Aldrich) were added, and phase-separated by centrifuged at 16,000xg for 15 minutes at 4oC. RNA was precipitated from the aqueous phase with ice-cold isopropanol and 15µg of glycogen (GlycoBlue™ Coprecipitant; Invitrogen) for 2 hours or overnight at 4oC. After centrifugation at 16,000xg for 30 minutes (at 4oC), the precipitated RNA pellets were washed with 75% ethanol, air-dried at room temperature and solubilized in 87.5µL of DEPC-treated water (Invitrogen). Residual genomic DNA was subsequently removed by in-solution digestion with 7U of RNase-free DNase I (Qiagen). The RNA was then cleaned up by a second phenol-chloroform extraction step, and finally solubilized in 15µL DEPC-treated water. Absence of genomic DNA was determined by 35 cycles of RT-qPCR, using the skeleton binding protein 1 (*SBP1*) as target gene. If Ct values were less than 32, DNase digestion and re-extraction was repeated. The RNA was either used immediately or stored at -80oC.

### 4.7 Estimation of primer efficiency

Primers used for 3D7 *var* gene expression analysis were selected from previous studies [53, 3]. All primers were synthesized by Eurofins Genomics at 0.01 µmole with HPSF purification. *P. falciparum* 3D7 genomic DNA was serially diluted over 5-log concentrations and applied in a qPCR assay to determine primer amplification efficiency, using the SensiFAST™ SYBR No-Rox kit (Bioline) and primers at 300nM concentration. PCR was run with a LightCycler® 480 System (Roche). A two-step PCR was applied, with initial denaturation at 95oC for 3 minutes, followed by 40 cycles of annealing and extension at 62oC ramping at 4.8o C/s. A melting curve step was included to ascertain the specificity of the primers. Only primers with efficiencies between 1.8 and 2.2 were used for further analyses.

### 4.8 Real time (RT)-qPCR

cDNA was synthesized with the PrimeScript™ RT reagent Kit (Takara) using a combination of random hexamers (100µM) and oligo dT (50µM) primers in 20µL reaction volumes. cDNA was used in qPCR for the quantification of *var* genes in each sample. RT-qPCR assays were run with the SensiFAST™ SYBR No-Rox kit (Bioline) and primers at 300nM concentration on a LightCycler® 480 System (Roche). A two-step PCR was used, with initial denaturation at 95oC for 3 minutes followed by 40 cycles of annealing and extension at 62oC with 4.8oC/s ramp. Each assay included a 2.5µg genomic DNA positive control and no template (water) negative control. A melting curve step was included to ascertain the specificity of the primers. Four Plasmodium genes; *SBP1* (PF3D7_0501300), fructose-bisphosphate aldolase (PF3D7_1444800), arginyl-tRNA synthetase (PF3D7_1218600) and seryl tRNA synthetase (PF3D7_0717700) were included as housekeeping genes (references). Normalisation and calibration were done as previously described [3]. In brief, *SBP1* was used as the normalizer while 2.5ng genomic DNA was used for calibration. Relative quantification was calculated using 2*^−^*^ΔΔ^*^CT^* taking into consideration the individual primer amplification efficiencies [44].

### 4.9 Estimating infection characteristics

For comparing infection progression in-terms of growth rate, we calculated the PMR using a piece-wise log-linear model with latency as an additional parameter, given by:

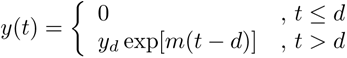

where *d* is the latency period, i.e. the time during which parasitaemia is undetectable, and *m* is the intrinsic growth rate of the parasite in each individual, *y* is the measured parasitaemia level, and *y_d_* is a small undetectable parasitaemia value. In the model we approximate to zero the undetectable parasitaemia during latency and consider that after latency the parasitaemia grows exponentially.

### 4.10 Estimating *var* gene expression changes

*var* gene expression probabilities were estimated as frequencies, i.e. ratios of the number of specific reads to the total number of reads. All probabilities less than a cut-off of 2% of the total *var* expression were considered as vanishing. Because each parasite expresses only one *var* gene at a time, the expression probability of a gene also represents the proportion of parasites expressing that gene.

To quantify the heterogeneity of the population of parasites in terms of quantity and type of *var* genes, we used the above expression probabilities to compute a Shannon diversity index as follows:

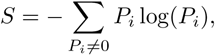

where *P_i_*represents the probability that the *var* gene *i* is expressed.

To model the change in *var* gene expression across time we considered a two state, continuous time^2^ Markov chain described by the diagram:

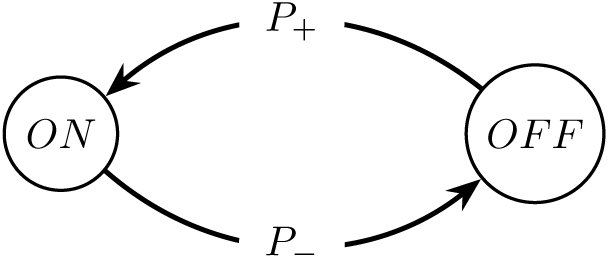

The two possible states for each variant are “ON” and “OFF” and *P*_+_ and *P_−_*are the probabilities of transition from one state to the other. The state “ON” means that the variant is expressed in a given parasite. The ’OFF’ state corresponds to both events in which the parasite has switched away to expressing another gene, as well as the parasite expressing the same gene was recognized and eliminated by the host immune system.

The probabilities of a gene being expressed at various times *t* + Δ*t* and at *t* are related by the following expression:

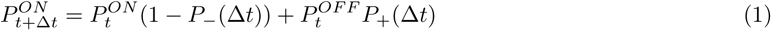

where *P_−_*(Δ*t*), *P*_+_(Δ*t*) are the finite time transition probabilities from OFF to ON and from ON to OFF, respectively. Using Eq.1, we can find bounds for the finite time transition probabilities *P_−_*(Δ*t*) and *P*_+_(Δ*t*). The probability bounds for *P_−_* can be given as:

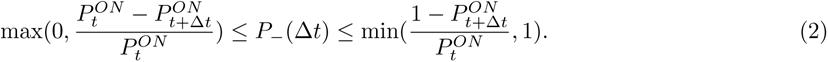

Similarly, the finite time transition probability from OFF to ON state satisfies:

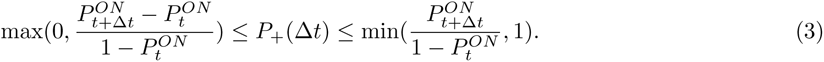

The finite time transition probabilities thus calculated depend on the time interval Δ*t*. To quantify the transition probabilities independently of the time interval, we estimate the instantaneous transition rate (per unit time) *p_−_*from ON to OFF, for each gene. Neglecting switching to events, instantaneous transition rate is related to the finite-time transition probability, as described by the formula:

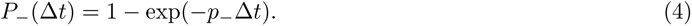

The finite time transition probability along a time Δ*t* equal to the generation time *T_g_*is equivalent to the rates derived previously in [27], using the discrete difference equations. Indeed, let us suppose that the gene only changes from ON to OFF. Then, the changes of the proportion of parasites expressing a variant *P_t_*over successive generations, starting with a monoclonal population *P*_0_ = 1 to represent the experimental conditions from [27], are described as:

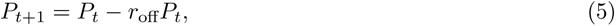

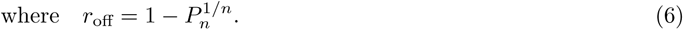

Comparing Eq. (1) with *P*_+_ = 0 to Eq. (5) we find the equivalence between *P_−_*(*T_g_*) and the discrete model rate

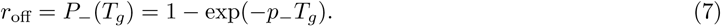

To summarize, in this paper we use both instantaneous transition rates (measured in d*^−^*^1^) and finite time transition probabilities (measured in % per generation) to estimate rates of changes of *var* gene expression. The above definitions are general and apply to both *in vivo* turnover and to *in vitro* switching, even if the mechanisms of gene population change are different. The definition of rates used in previous works [27], based on discrete Markov chains models and concerning *in-vitro* switching, is the same as our definition of finite time transition probabilities.

### 4.11 Modeling *in vitro* data

The analysed data consists of ten time series with up to six time points. The time series is multi-dimensional, as for each time point one has the probability of expression of each of the 61 *var* genes.

First, the high-dimensional traces were projected onto a 3D space using Principal Component Analysis.

This can be modelled as a continuous time, four-state Markov chain. The number of states in the model is obtained by using the following principles: i) each state corresponds to genes from the same group, ii) var gene groups such as *E* and *BA* have very low probabilities at late times for all sero-low samples and most sero-high samples (see Figure 5 g,h) and are therefore excluded from the analysis.

The *var* genes dynamics is described by a continuous time, four-state Markov chain model. The master equation for this model reads:

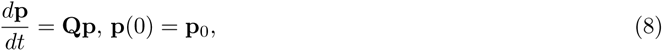

where **p**(*t*) = (*p*_1_(*t*)*, p*_2_(*t*)*, p*_3_(*t*)*, p*_4_(*t*))*^T^* and **p**_0_ are the time dependent and initial (*t* = 0) probabilities of the four states (in order, *var* gene groups A, B, C, BC), respectively, **Q** is the adjoint transition-rate matrix (also named adjoint infinitesimal generator matrix), satisfying *Q_ii_* = *−*Σ*_j≠i_ Q_ji_* (zero sum columnwise).

In this model, each state is a group of *var* genes. The element *Q_ji_, j* ≠ *i* represents the instantaneous transition rate from a state *i* to the state *j*. The instantaneous transition rates are estimated by optimisation and correspond to the minimum of the objective function

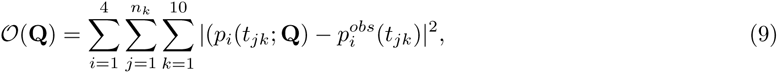

where *p_i_*(*t_ij_*; **Q**) are solutions of (8) with initial conditions **p**_0_ = **p***^obs^*(0) and *p^obs^*(*t_jk_*) are measured expression probabilities; the index 1 *≤ k ≤* 10 designates the sample and *n_k_≤* 6 is the number of time points for the sample *k*.

The instantaneous rates to switch away from each state are given by

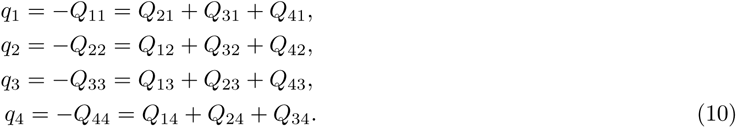

The finite time, per generation, probabilities to switch away can be computed from the diagonal elements of the matrix exp(*T_g_***Q**), where *T_g_* is the generation time, namely

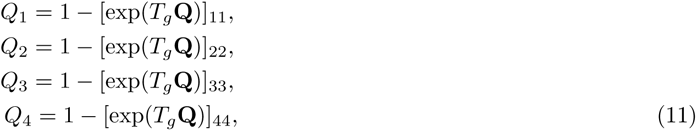

where exp(*∗*) represents the matrix exponential.

The inter-individual similarity of the large time *var* gene transcription profiles was tested using an AUC criterion. For an arbitrarily chosen late time sample the *var* genes were sorted in decreasing order of their expression values. Then a normalized rank was assigned to each *var* gene representing the rank in the particular sample divided by the total number of *var* genes. For all genes that have ranks smaller than a given one and for all samples, we computed the fraction of common genes that is the proportion of highly expressed *var* genes that are common to all late time samples. The AUC index was defined as the area comprised between the diagonal (fraction of common genes = normalized rank) and the fraction of common genes vs. normalised rank curve, low AUC meaning large similarity.

### 4.12 Protein microarray

We developed a custom microarray featuring extensive coverage of the PfEMP1 domains in the reference genome 3D7 as previously described [60] as well as 79 protein fragments from PfEMP1s from the IT4, HB3, and DD2 reference strains, as well as PfEMP1s sequenced from field isolates. PfEMP1 fragments were typically expressed as consecutive constitutive domains [47], as we have previously done with the reference genome 3D7 [60]. Intracellular acidic terminal segments (ATS) of PfEMP1s were expressed as stand-alone fragments. The microarray also included additional antigen malaria proteins, including the 3D7 variants of apical membrane antigen 1, circumsporozoite protein, and merozoite surface protein-1. Three concentrations of tetanus toxin were also included as positive controls. Construction of microarrays has been previously described elsewhere [**?** 57, 6]. The microarray was probed with plasma from study participants as previously described and then scanned [71]. Fluorescence intensity was defined as the raw signal intensity corrected by global median scaling for no-DNA negative controls.

### 4.13 Protein microarray data analysis

All statistical analyses were carried out in python version 3.9. MFI-bkg values smaller than or equal to zero, were replaced with the average value of blank responses (in this case with a value of 2 so that the *log*2*FC* value can be computed) and log-transformed. Sero-recognition threshold for all domains was determined by the median + 2 S.D. response levels in naive North-American individuals (n=10). Each domain on the array corresponds to a domain represented on the PfEMP1 protein encoded by the *var* genes. The random forest regressors and classifiers to predict relationships between immune response and infection outcome were implemented using scikit-learn 1.1.2. For data validation, we used responses against Tetanus antigens as control across naive and semi-immune individuals. The classification of the volunteers for immune response was retained as per the method described in the Epidemiological study design, based on another study, and later re-confirmed by several of our analyses. A PfEMP1 was considered recognised in an individual even if only one domain corresponding to the protein was recognised (in the event of multiple domains present per protein). The prediction of volunteer sub-groups was carried out based on immune responses using a random-forest classifier with Bootstrapping and Grid-Search to obtain optimal parameters for prediction of volunteers and a feature importance score was calculated to distinguish domain types useful in prediction the outcome of infection. We also calculated the change in breadth of response; as a cumulative total of domains recognised at different time-points in an individual, as well as the fold change in responses to each antigenic domain, between the first and the last time-point during the study.

### 4.14 *Var* expression and anti-PfEMP1 antibodies

A discretization method was used to compare inter-PfEMP1 responses throughout the CHMI. In this method, we converted the quantitative Fluorescence Intensity values per PfEMP1 to discrete values across all individuals, using quantile based classification. To discretize the domains, we used the following scheme: For each domain, we had a distribution of data points from samples defined as: *x*_1_*, x*_2_*, x*_3_*, …, x_n_*, where *n* is the number of individual samples per PfEMP1. We discretized the dataset into *k* + 1 bins using quantiles, where the quantiles are represented by *q*_1_*, q*_2_*, …, q_k_*. First, the quantile values *q*_1_*, q*_2_*, …, q_k_* were calculated based on the chosen number of categories. These quantiles divide the data into k+1 intervals. Then each data point *x_i_* was assigned to a specific interval based on its value. For instance, if *x_i_* falls in the interval (*q_j−_*_1_*, q_j_*], it is assigned to the j-th bin. Mathematically, this function can be represented as:

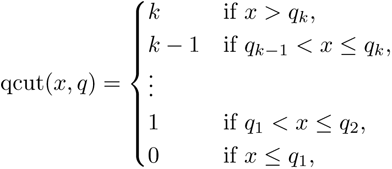

where *x* is a data point from the dataset, *q* = (*q*_1_*, q*_2_*, …, q_k_*), *k* is the number of quantiles.

We then grouped domains based on the PfEMP1 that they correspond to, and to each PfEMP1 group we associate the maximum domain response. Finally, only the PfEMP1s for which there was data available for both qPCR expression as well immune response were selected for the analysis.

## Data Availability

All data produced are available online at Zenodo

https://zenodo.org/records/10432857

## Acknowledgments

We thank Isaie Reuling for helping setting up the CHMI. This work was funded by grants from the joint MRC/LSHTM fellowship, the French National Research Agency (18-CE15-0009-01), Méditérannée Infection (InfectioPole Sud), ATIP-Avenir, Fonds Médical de la Recherche (FRM).

## Supplementary figures

**Figure S1:**
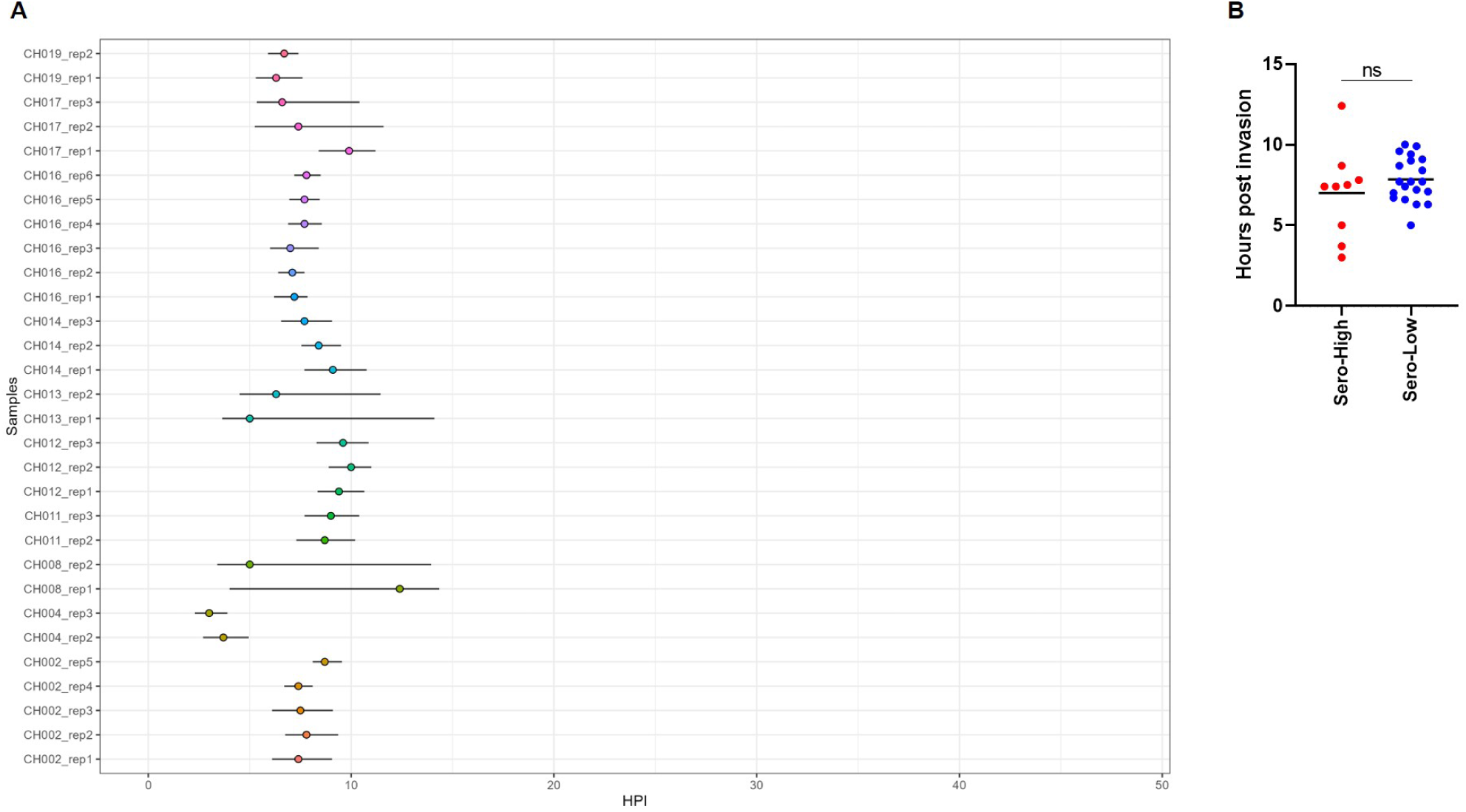
Developmental age of parasites by maximum likelihood estimated based on the captured transcriptomes *in vivo.* (A) All isolates were estimated to be ring-stages. (B) No significant difference between *sero-low* and *sero-high* derived isolates.

**Figure S2:**
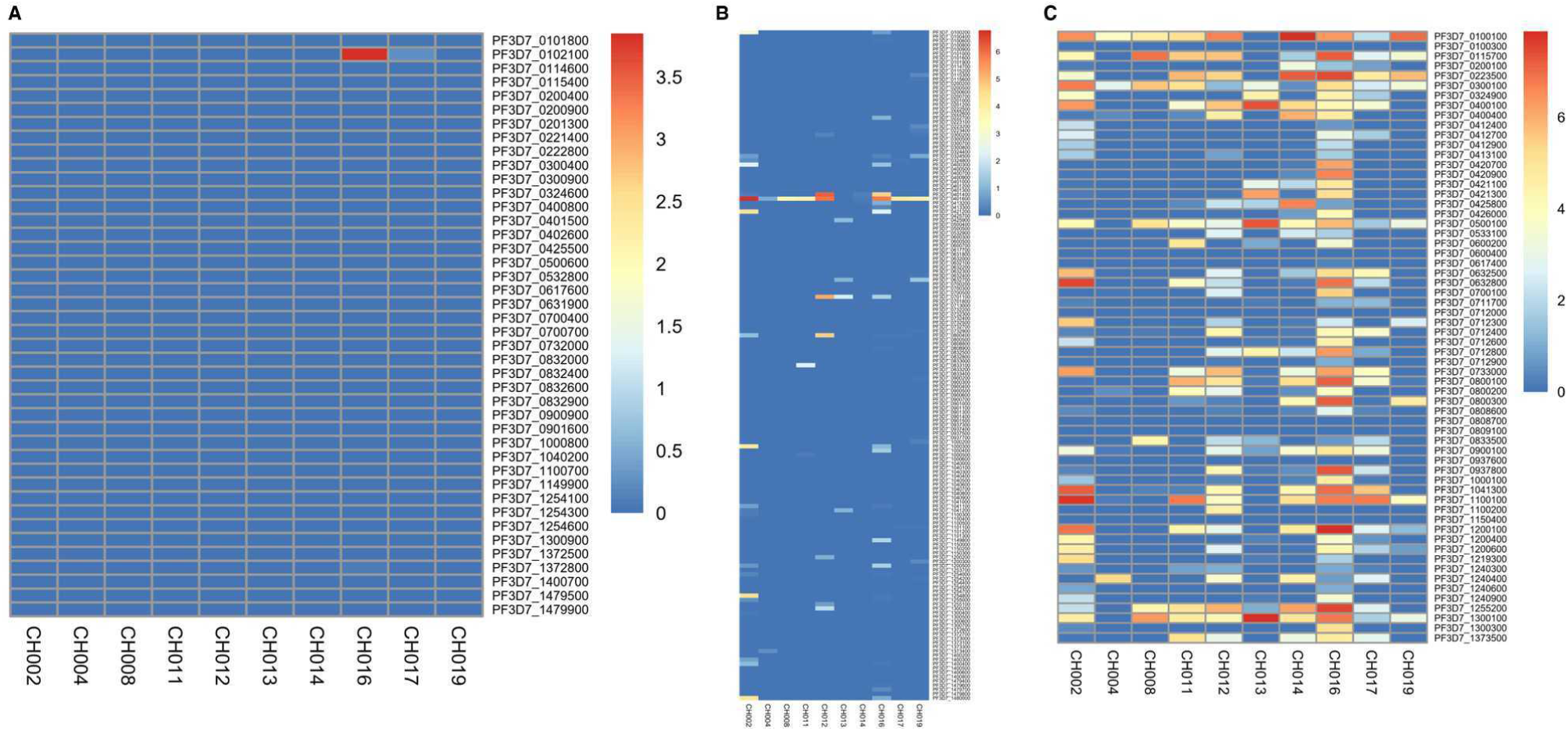
*In vivo* expression levels of variant surface antigens (VSA) from bulk RNA-seq.:Heatmap shows Log2FPKM values in each isolate. (A) *stevor* genes., (B) *rif* genes. (C) *var* genes.

**Figure S3:**
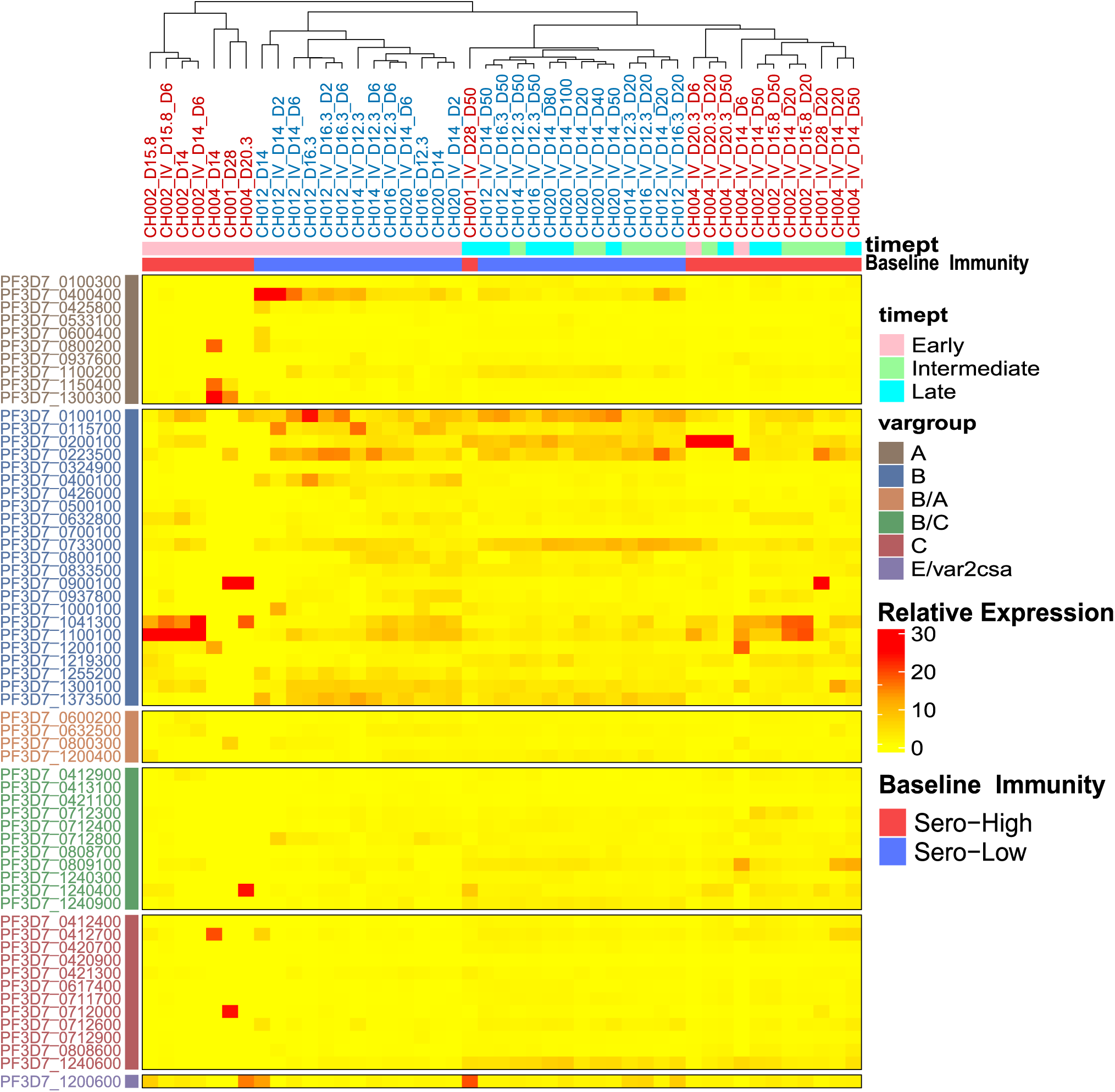
*In vitro var* expression landscape across time for ten isolates derived from *sero-low* and *sero-high* individuals. The heatmap is hierarchically clustered for relative expression of *var* genes at timepoints 6, 20 and 50 days *in vitro* after the *in vivo* infection timepoint. For CH020 (*sero-low* individual) there was an additional timepoint at 100 days post *in vivo*. Relative expression of each gene is shown from *low* to *high*, between 0% (yellow) to 30% (red). *Var* genes were grouped based on upstream sequence as A, B, C,E or intermediate groups B/A and B/C. Individuals were clustered based on expression and stratified by infection outcome.

**Figure S4:**
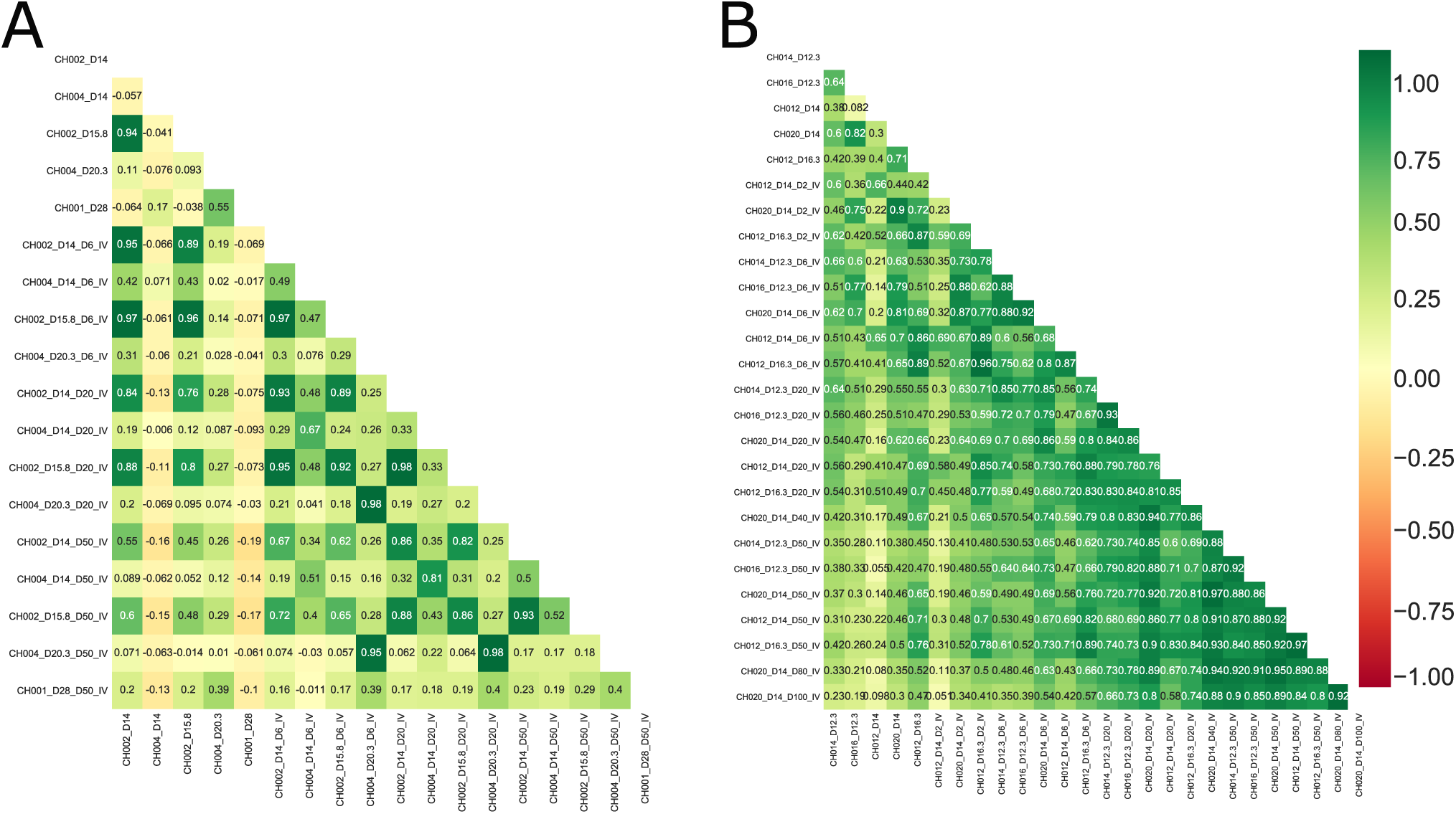
Pearson correlation between the *var* expression times series of A) *sero-high* individuals during *in vitro* culture and B) sero-low individuals. The color gradient is ordered from red (least) to green (highest).

**Figure S5:**
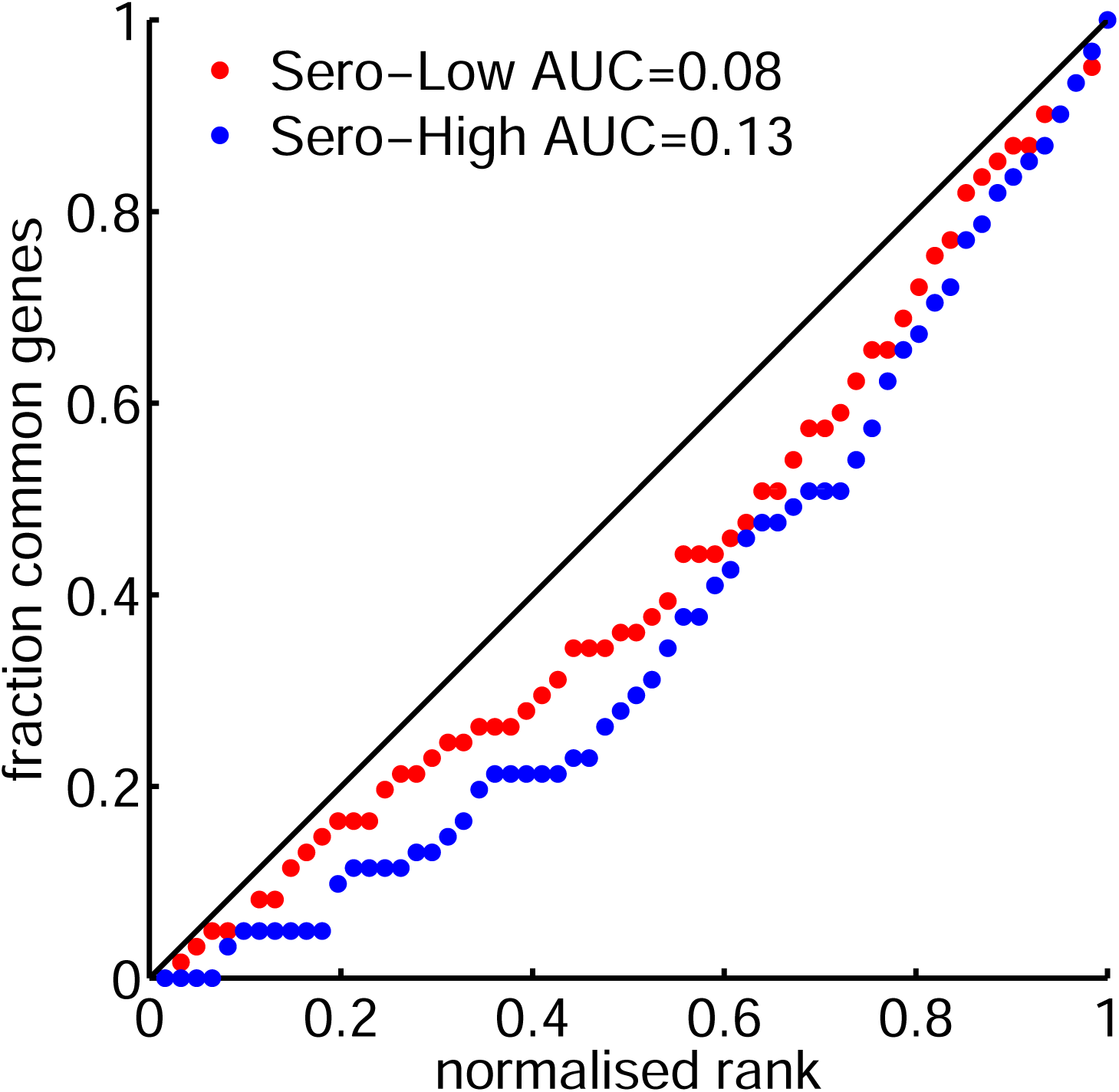
AUC method for testing the similarity of *var* genes distribution in all late time samples *in vitro*. The normalised rank represent the rank divided by total number of *var* genes after sorting them with respect to expression value in decreasing order. The fraction of common genes is the proportion of *var* genes that have ranks smaller than a given one in all late time samples. The diagonal represents the perfect similarity. The AUC index is defined as the area comprised between the diagonal and the curve, lower AUC meaning larger similarity.

**Figure S6:**
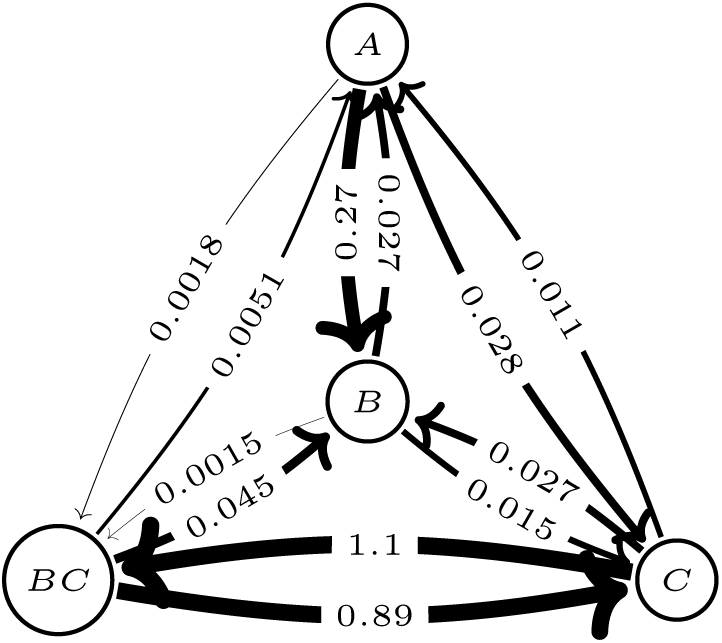
Transition graph and instantaneous switching rates of the four states *var* genes model resulting from fitting the model to the *in vitro* data. The gene groups *E* and *BA* where not included in this model because they reach rapidly very low steady state probabilities (see Figure 5 g,h).

**Figure S7:**
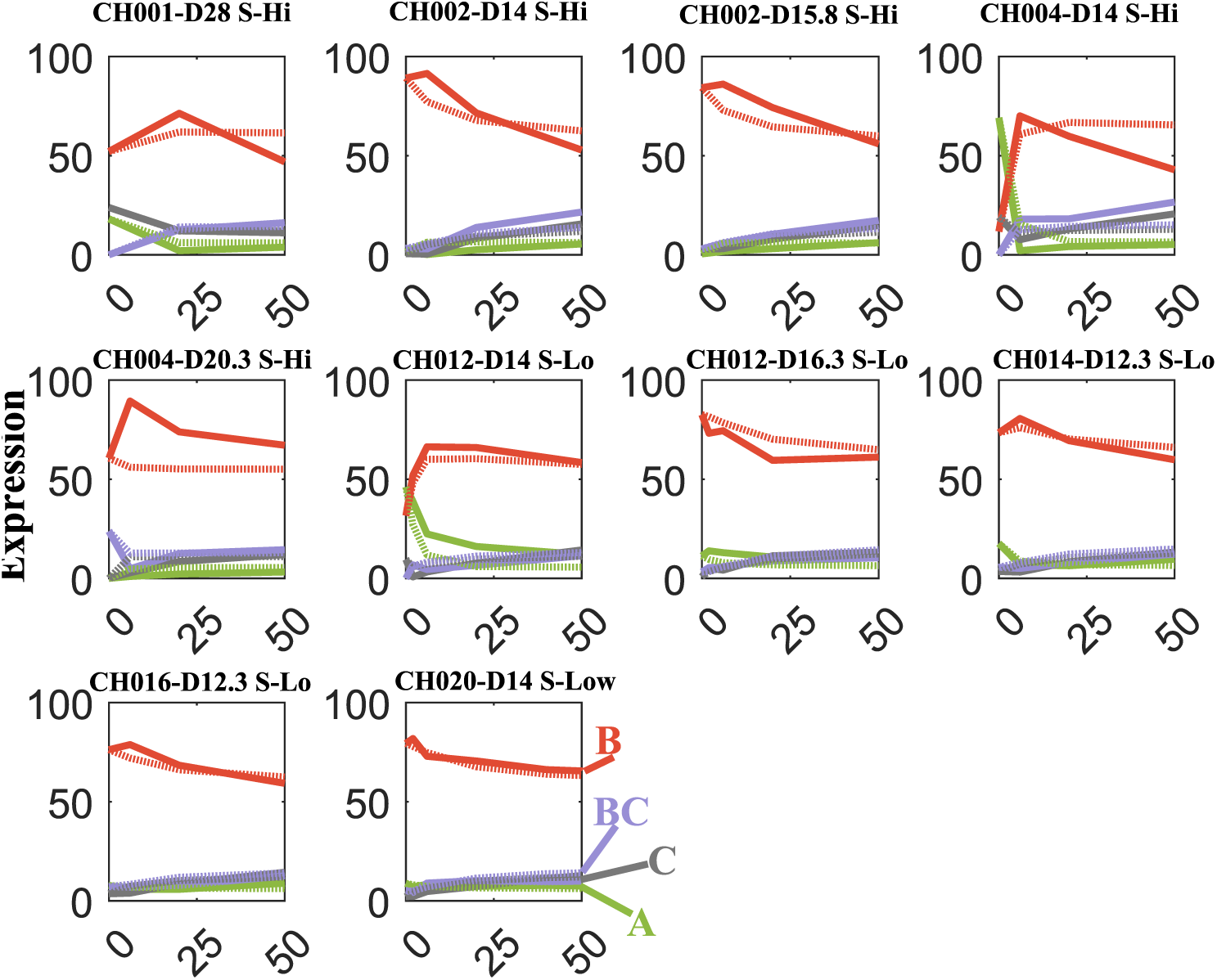
Fitting the four-state model to the *in vitro* expression data. Dotted lines are the model predictions.

**Figure S8:**
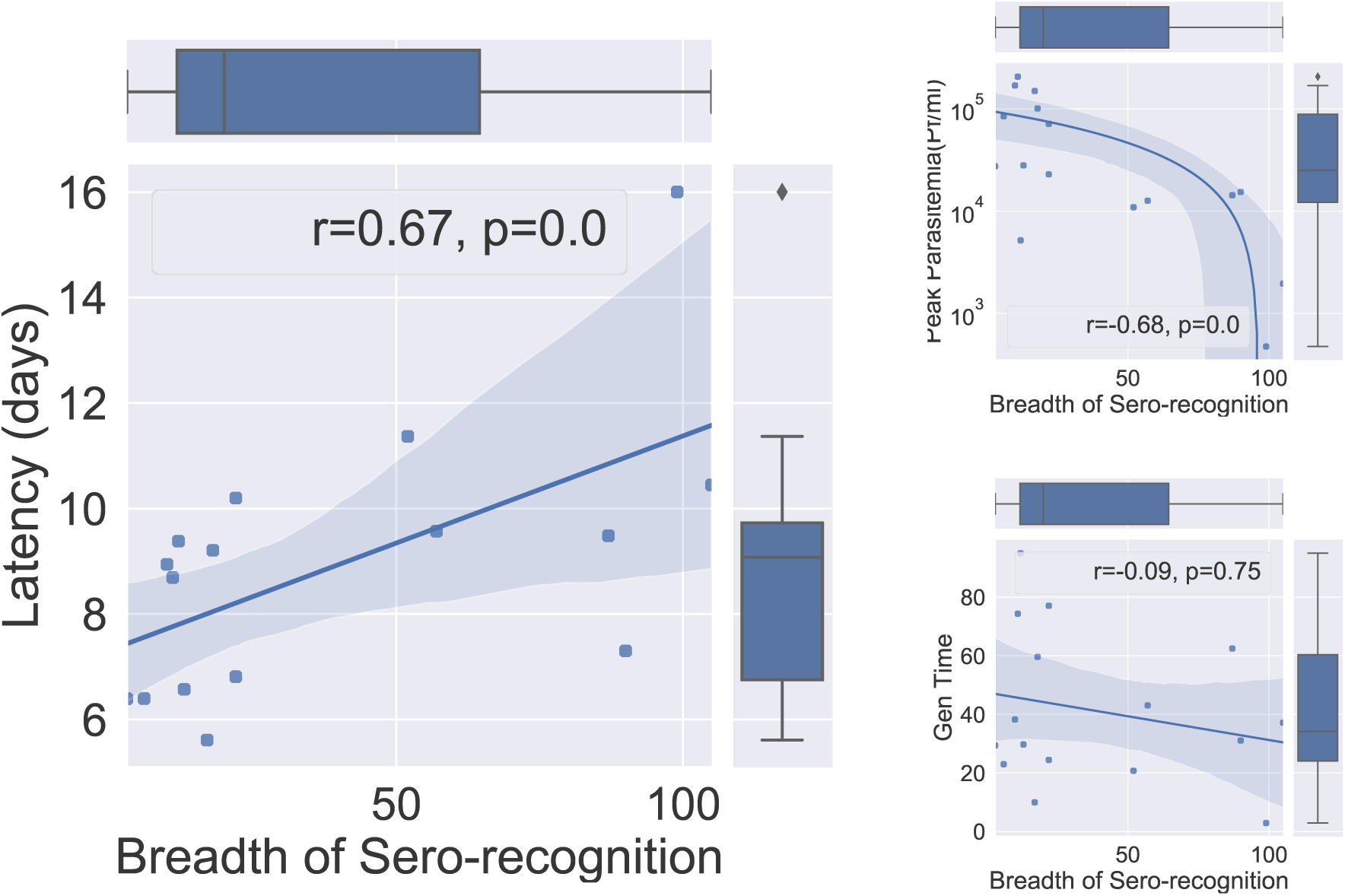
Pre-infection breadth of immune response and parasite growth dynamics. A) Spearmann Rank correlation between the number of 3D7 PfEMP1 domains recognized by the host at the beginning of the infection and the latency period (days until patent parasitaemia). B) Sero-recognition at day 0 and peak of parasitaemia: The log-peak of parasitaemia (Pf/ml) correlation with respect to the number of sero-recognized domains at the start of infection in an individual. C) Spearmann Correlation plot for parasite multiplication rate vs breadth of sero-recognition in each individual.

## Footnotes

1 the first and second part of the sample symbol indicates the volunteer and the the day after infection when the isolate was drawn, respectively

2 Even if one parasite switches the variant only at discrete times, multiples of the generation time, a continuous-time model is appropriate for modeling a population of non-synchronized parasites.

